# BCL11A intellectual developmental disorder: defining the clinical spectrum and genotype-phenotype correlations

**DOI:** 10.1101/2021.09.06.21262776

**Authors:** Angela Peron, Felice D’Arco, Kimberly A. Aldinger, Constance Smith-Hicks, Christiane Zweier, Gyri A. Gradek, Kimberley Bradbury, Andrea Accogli, Erica F. Andersen, Ping Yee Billie Au, Roberta Battini, Daniah Beleford, Lynne M. Bird, Arjan Bouman, Ange-Line Bruel, Øyvind Løvold Busk, Philippe M. Campeau, Valeria Capra, Colleen Carlston, Jenny Carmichael, Anna Chassevent, Jill Clayton-Smith, Michael J Bamshad, Dawn L. Earl, Laurence Faivre, Christophe Philippe, Patrick Ferrerira, Luitgard Graul-Neumann, Mary J. Green, Darrah Haffner, Parthiv Haldipur, Suhair Hanna, Gunnar Houge, Jane Hurst, Cornelia Kraus, Birgit Elisabeth Kristiansen, James Lespinasse, Karen J. Low, Sally Ann Lynch, Sofia Maia, Rong Mao, Ruta Marcinkute, Catherine Melver, Kimberly McDonald, Tara Montgomery, Manuela Morleo, Constance Motter, Amanda S. Openshaw, Janice Cox Palumbos, Aditi Shah Parikh, Richard Person, Megha Desai, Juliette Piard, Rolph Pfundt, Marcello Scala, Margaux Serey-Gaut, Anne Slavotinek, Mohnish Suri, Claire Turner, Tatiana Tvrdik, Karin Weiss, Ingrid M. Wentzensen, Marcella Zollino, C4RCD Research Group, Telethon Undiagnosed Disease Program (TUDP), University of Washington Center for Mendelian Genomics (UW-CMG), Bert B.A. de Vries, Francois Guillemot, William B. Dobyns, David Viskochil, Cristina Dias

**Author notes:** **Corresponding authors:** Dr. Angela Peron, Dr. Cristina Dias. A complete list of members and their affiliations appears in the Supplementary Data (C4RCD Research Group, Telethon Undiagnosed Disease Program (TUDP), University of Washington Center for Mendelian Genomics (UW-CMG)).

## Abstract

**Purpose:** Heterozygous variants in *BCL11A* underlie an intellectual developmental disorder with persistence of fetal hemoglobin (BCL11A-IDD, a.k.a. Dias-Logan syndrome). We sought to delineate the genotypic and phenotypic spectrum of BCL11A-IDD.

**Methods:** We performed an in-depth analysis of 42 patients with BCL11A-IDD ascertained through a collaborative network of clinical and research colleagues. We also reviewed 33 additional affected individuals previously reported in the literature or available through public repositories with clinical information.

**Results:** Molecular and clinical data analysis of 75 patients with BCL11A-IDD identified 60 unique variants (30 frameshift, 7 missense, 6 splice-site, 17 stop-gain) and 8 unique CNVs (microdeletions involving *BCL11A* only). We redefined the most frequent manifestations of the condition: intellectual disability, hypotonia, behavioral abnormalities, postnatal microcephaly and autism spectrum disorder. Two thirds of patients have brain MRI abnormalities, and we identified a recurrent posterior fossa phenotype of vermian hypoplasia and/or small brainstem. Truncating *BCL11A* variants, particularly those affecting the long (BCL11A-L) and extra-long (-XL) isoforms, sparing the short (-S) isoform, were associated with increased severity.

**Conclusions:** We expand the clinical delineation of BCL11A-IDD and identify a potential isoform-specific genotype-phenotype correlation. We show that BCL11A-IDD is associated with posterior fossa anomalies and highlight the differences between BCL11A-IDD and 2p16.1p15 microdeletion syndrome.

## Introduction

BCL11A-Related Intellectual Developmental Disorder (BCL11A-IDD, also known as intellectual developmental condition with persistence of fetal hemoglobin [OMIM # 617101] or Dias-Logan syndrome) is an autosomal dominant disorder caused by heterozygous pathogenic variants in *BCL11A*^1^. *BCL11A* encodes a Krüppel-like sequence-specific C2H2 zinc finger transcription factor (BCL11A, a.k.a. CTIP1). It has been extensively investigated for its role in hematopoiesis and the fetal-to-adult hemoglobin switch via transcriptional repression ^2^, whereby the BCL11A internal zinc finger domains selectively bind to γ-globin promoter motifs^3^. BCL11A’s relevance in neurodevelopment has come to light in recent years. It is highly expressed in the developing mammalian brain^4,5^ and regulates multiple developmental processes, including subtype identity in deep-layer projection neurons^6^, differentiation and thalamocortical integration of layer IV projection neurons^7^, and late differentiation, cell-polarity switch and radial migration of upper-layer projection neurons^8^. It is also required for correct sensory circuit formation of the dorsal spinal cord. We have previously demonstrated that haploinsufficiency of BCL11A leads to impaired memory and social behavior in mice, as well as regionalized reduction of brain size^5^.

A pivotal demonstration of BCL11A’s importance to human brain development has been the identification of individuals with neurodevelopmental phenotypes associated with heterozygous mutation or copy number loss of *BCL11A*^5,9-11^. BCL11A-IDD has been identified in individuals with heterozygous loss-of-function variants^5,12-16^, as well as N-terminus missense variants acting as hypomorphic alleles, reducing BCL11A transcriptional activity and subcellular localization^5^. Despite these reports, a detailed understanding of the phenotypic spectrum of BCL11A-IDD is lacking, as is the effect of genotypic variation on disease features, severity and prognosis (genotype-phenotype correlation). Moreover, multiple isoforms have partial or complete annotation for human and mouse *BCL11A* (see www.ensembl.org), where possible tissue and developmental period specificity of isoform expression pose further challenges. There is experimental evidence for the expression of three main BCL11A isoforms (extra-long (XL), long (L), and short (S)) in developing human brain ^17,18^, herein designated BCL11A-XL (835 aa), BCL11A-L (773 aa), and BCL11A-S (243 aa).

In light of the above, we aimed to further define the phenotypic spectrum of BCL11A-IDD and determine genotype-phenotype correlations by analyzing a large cohort of affected individuals. Here we provide detailed phenotypic data on 42 individuals with *BCL11A* variants consistent with BCL11A-IDD and review an additional 33 case reports. We demonstrate a broad spectrum of severity of neurodevelopmental phenotypes and mild dysmorphic features. We provide further evidence for previously under-appreciated phenotypes including hindbrain abnormalities and novel rare autonomic phenotypes. We evaluate the effect of variants in the context of different *BCL11A* isoforms and identify a putative isoform-specific genotype-phenotype effect.

## Materials and Methods

### Patients

Present cohort: Individuals with BCL11A-IDD were identified through collaborating clinicians’ direct clinical practice, the Deciphering Developmental Disorders Study,^10^ GeneMatcher^19^ and by contact with submitters of molecularly diagnosed individuals in publicly available databases: Decipher^20^ and ClinVar.^21^ For the latter, submitting diagnostic laboratories kindly facilitated contact with referring clinicians within the terms and conditions each provider so permitted. Patients previously reported by co-authors are included in the present cohort with updated phenotypic information: P4,^22^ P8 and P24,^23^ P17, P30, P35, P36.^5^ All patients had been previously identified as having a likely pathogenic or pathogenic variant in *BCL11A*. Details of the prior genetic analyses performed are provided in Supplementary Data. Informed consent for participating in diagnostic studies was obtained by referring clinicians at local hospitals. Subjects identified and assessed through research studies were consented according to institutional review boards of local regulatory authorities (see Ethics Declaration). All necessary patient/participant consent has been obtained and the appropriate institutional forms have been archived.

Additional previously reported individuals were identified via literature search in PubMed (last search date May 9, 2021). The following search terms were used: “BCL11A”, “Dias-Logan syndrome”, “2p16.1 deletion”. For patients with microdeletions, those with copy number variants (CNVs) encompassing only *BCL11A* or *BCL11A* and non-coding genes were included for assessment. Those with larger genomic deletions encompassing additional protein-coding genes were excluded from this cohort and included only as a comparison group of selected phenotypes (last search date July 1, 2021). Individuals with additional likely pathogenic or pathogenic CNVs in addition to 2p15-2p16.1 were excluded.

Open access pathogenic and likely pathogenic variants (where the submitter was unavailable or unable to provide further information) were reviewed from publicly available databases DECIPHER and ClinVar (last search date April 17, 2021). Only variants where at least one phenotypic term was submitted were included.

### Phenotyping

Collaborating clinicians were asked to provide retrospectively collected clinical and molecular information regarding previously identified patients on a standardized questionnaire. Previously reported individuals enrolled in the BUILD clinical research study (see Ethics Declaration) were prospectively clinically reassessed. In addition to clinical features provided by the referring clinician, where photographs were available for analysis with patient/guardian consent, facial features were assessed independently by two clinical geneticists experienced in dysmorphology (A.P. and C.D.) and a final consensus facial phenotype was recorded.

Clinical manifestations were annotated using Human Phenotype Ontology (HPO) terms wherever possible^24^ (Supplementary Table S1). Facial features were reported using the nomenclature of the Elements of Morphology: Standard Terminology.^25^ Where growth parameters were recorded as raw numbers, age and sex adjusted standard deviations (SD) were calculated using the UKWHO reference in the childsds package version 0.7.6^26^ in R version 4.0.3. Percentiles/SD provided by the examining clinician were used where primary measurements were unavailable.

### Variant annotation

Given the discrepancy between BCL11A transcript assemblies for GRCh37 and GRCh38, variants with chromosomal positions annotated to GRCh37 were mapped to GRCh38 using Assembly Converter (Ensembl release 103 - February 2021). All variants (SNVs and CNVs) were annotated to RefSeq transcripts using Variant Effect Predictor with genome build GRCh38.p13 (Ensembl). CNVs were visualized as a custom track in Ensembl (GRCh38.p13). *BCL11A* has multiple annotated transcripts; for the purposes of this manuscript we selected 3 transcript isoforms with evidence of expression in brain tissue^17^ and use the following nomenclature: transcripts *BCL11A*-XL (NM_022893.4, MANE transcript), *BCL11A*-L (NM_018014.4) and *BCL11A*-S (NM_138559.2), encoding the following proteins, respectively, BCL11A-XL (NP_075044.2, 835 aa), BCL11A-L (NP_060484.2, 773 aa), and BCL11A-S (NP_612569.1, 243 aa).

Variants were classified based on predicted effects on the different isoforms and on predictions of nonsense mediated decay escape (NMDe).^27^ *PTVa*, premature termination codon (PTC) in all 3 isoforms: *PTVa1*, possible escape or decreased efficiency of NMD; *PTVa2*, likely NMD in all 3 isoforms. *PTVb*, PTC affects only BCL11A-XL and BCL11A-L (downstream of BCL11A-S naturally occurring termination codon): *PTVb1*, PTC or frameshift upstream of all zinc finger regions; PTVb2, PTC or frameshift upstream of C-terminus zinc fingers. *MISS*: missense variants. *SPL*: splice variants predicted to alter the reading frame. *CNV*: copy number variations (microdeletions).

### Fetal forebrain and cerebellum

Human fetal hindbrain 16pcw (post-conception weeks) and 18pcw were provided by the Joint MRC/Wellcome Trust (grant# MR/R006237/1) Human Developmental Biology Resource (www.hdbr.org). The HDBR has ethical approval (REC 18/NE/0290). Human fetal hindbrain 12pcw was obtained from the Birth Defects Research Laboratory (BDRL) at the University of Washington, Seattle, WA with ethics board approval and maternal written consent. This study was performed in accordance with ethical and legal guidelines of Seattle Children’s Hospital Institutional Review Board. Immunohistochemistry was performed using standard methods detailed in the Supplementary Data.

### Neuroradiology analysis

Where images were available, retrospectively acquired brain MRI scans were analyzed by an experienced pediatric neuroradiologist (F.D’A.), provided with a known diagnosis of BCL11A-IDD but blinded to mutation. MRI scans of 13 patients were analyzed for presence of abnormalities in the brain (and spine in a subset). The abnormalities in the posterior fossa were assessed qualitatively and using quantitative measurements when DICOM images were available: the vermian height, vermian antero-posterior diameter, antero-posterior midbrain-pons junction and antero-posterior mid-pons diameter were measured on the sagittal midline and compared with normal values for age-matched controls available in literature ^28^. Values were considered abnormal when below the 3rd percentile. The pons was considered abnormal when the cranio-caudal ratio with midbrain and/or medulla oblongata was less than 1.5:1.^29^

### Statistical analysis

The frequencies of each clinical manifestation were calculated using as the denominator the number of individuals for whom clear evidence of presence/absence of each manifestation was available (i.e. patients where the data were not available were excluded from the calculation). Categorical variables were described as n (%), and continuous variables were described as median and ranges. Statistical tests were performed using the Graphpad Prism software v.8.0.1. Graphical representations were generated using Graphpad Prism and R v.4.0.3 (ggplot2 v.3.3.3, circlize v.0.4.13). Unless otherwise indicated, for comparison of frequencies between groups, Fisher’s exact test was used, at a significance threshold of *p<0*.*05*. For comparison over timepoints, 2-way ANOVA was applied.

## Results

### Cohort overview

We present 42 individuals with heterozygous variants in *BCL11A* [18 females (43%) and 24 males (57%)] classified as pathogenic or likely pathogenic. Of these, 37 are previously unreported; 5 have been reported previously,^5,22^ but are presented here with detailed update of the phenotype. Median age at molecular diagnosis was 7 years (range 1-19 y). Median age at last assessment was 9 years (range 2-23 y) (Supplementary Table S1). We also analyzed data from 17 previously published patients^5,12-16^ and from 16 records deposited in publicly available databases, for a total of 75 patients with BCL11A-IDD. Median age at last assessment of the previously published individuals was 7^+5/12^ years (range 3-15 y). Median age at last assessment of the patients in publicly available databases was 5^+11/12^ years (range 3^+7/12^-7^+5/12^ y). Table 1 shows the main clinical manifestations of each subgroup and of the combined cohort. Details on each individual can be found in Supplementary Table S1. To account for potential biases in clinical characterization, we present the results of the present cohort (n=42, unpublished or updated patients) and of the “combined cohort” (n=75, present cohort together with previously reported – in the literature and open access databases) separately.

**Table 1:**
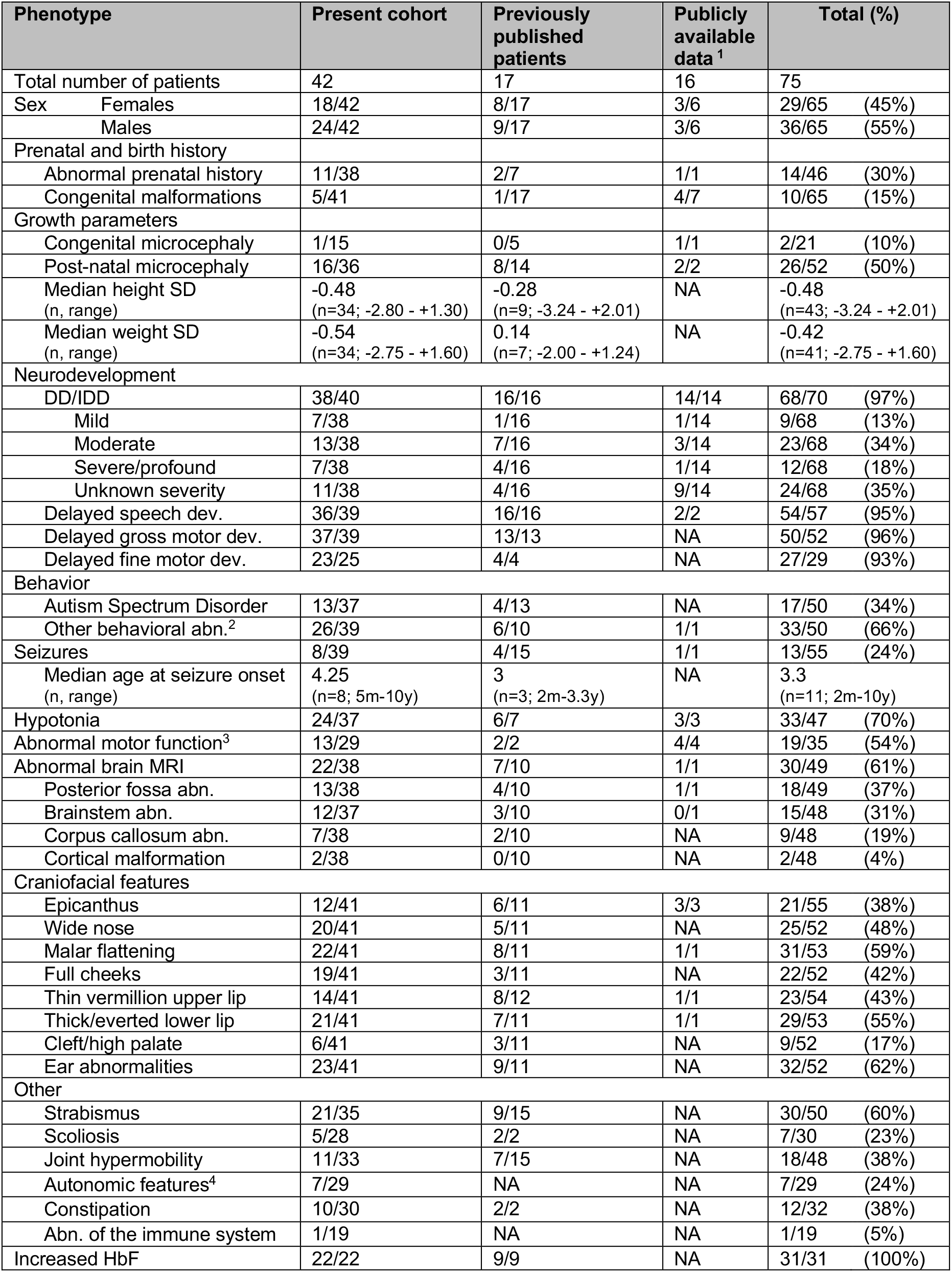
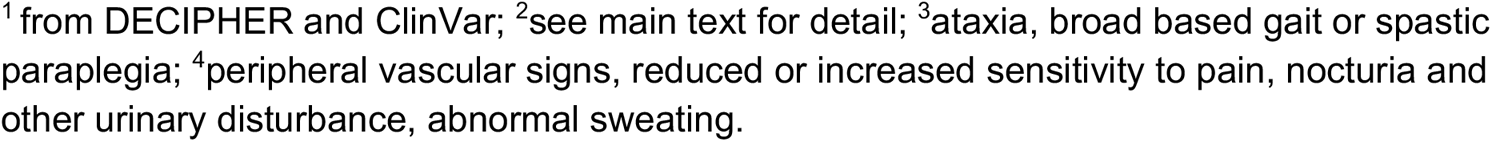
Demographic and clinical information about individuals with pathogenic and likely pathogenic variants in *BCL11A*. HPO terms for each clinical manifestation can be found in Supplementary Table S1. NA: Not Available; SD: Standard Deviation; DD: developmental delays; IDD: intellectual developmental disorder; dev.: development; abn.: abnormalities; m, months; y, years MRI: magnetic resonance imaging; HbF: fetal hemoglobin;

| Phenotype | Present cohort | Previously published patients | Publicly available data<sup>1</sup> | Total (%) |
| --- | --- | --- | --- | --- |
| Total number of patients | 42 | 17 | 16 | 75 |
| Sex Females | 18/42 | 8/17 | 3/6 | 29/65 (45%) |
| Males | 24/42 | 9/17 | 3/6 | 36/65 (55%) |
| Prenatal and birth history |  |  |  |  |
| Abnormal prenatal history | 11/38 | 2/7 | 1/1 | 14/46 (30%) |
| Congenital malformations | 5/41 | 1/17 | 4/7 | 10/65 (15%) |
| Growth parameters |  |  |  |  |
| Congenital microcephaly | 1/15 | 0/5 | 1/1 | 2/21 (10%) |
| Post-natal microcephaly | 16/36 | 8/14 | 2/2 | 26/52 (50%) |
| Median height SD (n, range) | -0.48 (n=34; -2.80 - +1.30) | -0.28 (n=9; -3.24 - +2.01) | NA | -0.48 (n=43; -3.24 - +2.01) |
| Median weight SD (n, range) | -0.54 (n=34; -2.75 - +1.60) | 0.14 (n=7; -2.00 - +1.24) | NA | -0.42 (n=41; -2.75 - +1.60) |
| Neurodevelopment |  |  |  |  |
| DD/IDD | 38/40 | 16/16 | 14/14 | 68/70 (97%) |
| Mild | 7/38 | 1/16 | 1/14 | 9/68 (13%) |
| Moderate | 13/38 | 7/16 | 3/14 | 23/68 (34%) |
| Severe/profound | 7/38 | 4/16 | 1/14 | 12/68 (18%) |
| Unknown severity | 11/38 | 4/16 | 9/14 | 24/68 (35%) |
| Delayed speech dev. | 36/39 | 16/16 | 2/2 | 54/57 (95%) |
| Delayed gross motor dev. | 37/39 | 13/13 | NA | 50/52 (96%) |
| Delayed fine motor dev. | 23/25 | 4/4 | NA | 27/29 (93%) |
| Behavior |  |  |  |  |
| Autism Spectrum Disorder | 13/37 | 4/13 | NA | 17/50 (34%) |
| Other behavioral abn.<sup>2</sup> | 26/39 | 6/10 | 1/1 | 33/50 (66%) |
| Seizures | 8/39 | 4/15 | 1/1 | 13/55 (24%) |
| Median age at seizure onset (n, range) | 4.25 (n=8; 5m-10y) | 3 (n=3; 2m-3.3y) | NA | 3.3 (n=11; 2m-10y) |
| Hypotonia | 24/37 | 6/7 | 3/3 | 33/47 (70%) |
| Abnormal motor function<sup>3</sup> | 13/29 | 2/2 | 4/4 | 19/35 (54%) |
| Abnormal brain MRI | 22/38 | 7/10 | 1/1 | 30/49 (61%) |
| Posterior fossa abn. | 13/38 | 4/10 | 1/1 | 18/49 (37%) |
| Brainstem abn. | 12/37 | 3/10 | 0/1 | 15/48 (31%) |
| Corpus callosum abn. | 7/38 | 2/10 | NA | 9/48 (19%) |
| Cortical malformation | 2/38 | 0/10 | NA | 2/48 (4%) |
| Craniofacial features |  |  |  |  |
| Epicanthus | 12/41 | 6/11 | 3/3 | 21/55 (38%) |
| Wide nose | 20/41 | 5/11 | NA | 25/52 (48%) |
| Malar flattening | 22/41 | 8/11 | 1/1 | 31/53 (59%) |
| Full cheeks | 19/41 | 3/11 | NA | 22/52 (42%) |
| Thin vermilion upper lip | 14/41 | 8/12 | 1/1 | 23/54 (43%) |
| Thick/everted lower lip | 21/41 | 7/11 | 1/1 | 29/53 (55%) |
| Cleft/high palate | 6/41 | 3/11 | NA | 9/52 (17%) |
| Ear abnormalities | 23/41 | 9/11 | NA | 32/52 (62%) |
| Other |  |  |  |  |
| Strabismus | 21/35 | 9/15 | NA | 30/50 (60%) |
| Scoliosis | 5/28 | 2/2 | NA | 7/30 (23%) |
| Joint hypermobility | 11/33 | 7/15 | NA | 18/48 (38%) |
| Autonomic features<sup>4</sup> | 7/29 | NA | NA | 7/29 (24%) |
| Constipation | 10/30 | 2/2 | NA | 12/32 (38%) |
| Abn. of the immune system | 1/19 | NA | NA | 1/19 (5%) |
| Increased HbF | 22/22 | 9/9 | NA | 31/31 (100%) |

<sup>1</sup>from DECIPHER and ClinVar; <sup>2</sup>see main text for detail; <sup>3</sup>ataxia, broad based gait or spastic paraplegia; <sup>4</sup>peripheral vascular signs, reduced or increased sensitivity to pain, nocturia and other urinary disturbance, abnormal sweating.

### Mutational spectrum

The present cohort includes 31 patients with 26 unique protein truncating variants (PTV) [10 patients carry PTVa variants (9 unique variants), affecting all 3 isoforms, and 21 patients carry PTVb (17 unique variants), sparing BCL11A-S], 3 splice variants (SPL), 4 missense variants (MISS) (Figure 1, Supplementary Figure S1A, Supplementary Table S2) and 4 unique copy number variants (CNV) (Supplementary Figure S1B, Supplementary Tables S3). Only 5 variants are predicted to efficiently undergo nonsense mediated decay (NMD) in all 3 isoforms (PTVa2). All other variants are predicted to escape or have reduced efficiency of NMD via different mechanisms, leading to truncated proteins with or without changes to the open reading frame (Supplementary Table S2). Overall, the combined cohort includes 53 patients with 47 unique PTV variants (20 PTVa and 33 PTVb), 6 SPL, 7 MISS and 9 CNV (8 unique).

**Figure 1.**
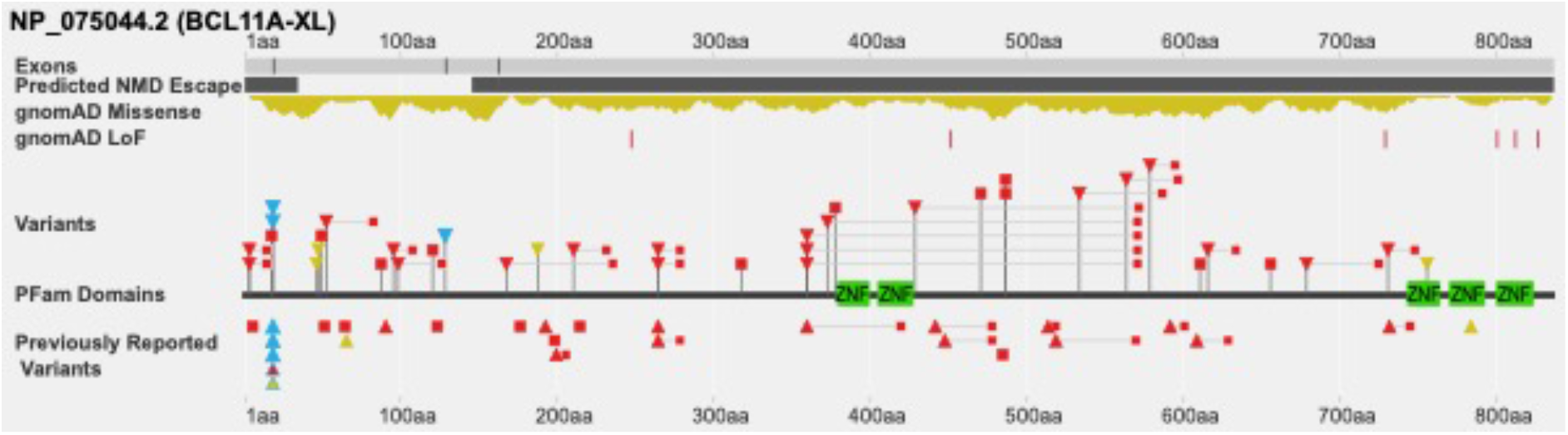
BCL11A genomic variants annotated to MANE isoform BCL11A-XL. Stop gain: red square; frameshift: red triangle, with downstream premature termination codon as small red square; missense variants: yellow triangle; splice variants: light blue triangle. Light blue outline on red triangle: frameshift and potential splice variant ClinVar_VCV000987092.1; light blue outline on yellow triangle: missense and potential splice variant ClinVar_VCV000987093.1 (see Supplementary Table S2). Green: C2H2 DNA binding zinc finger domains. Dark grey bars: regions of predicted NMD escape by “start proximal” and “last exon junction” mechanisms. This figure was generated in collaboration with the Decipher team (https://deciphergenomics.org).

Information on inheritance was available for 38 affected individuals of the present cohort. Thirty-one patients (82%) had *de novo* variants. In four patients from 2 families germline mosaicism was assumed, as unaffected parents had two affected children but tested negative on blood and/or buccal swab testing. Two affected individuals inherited the variant from an apparently unaffected parent, although formal neurocognitive assessment was not performed in them. In one affected individual, the variant was not maternally inherited (paternal DNA unavailable).

Previous genetic testing for each patient is summarized in Supplementary Table S1. Six individuals from the present cohort had additional CNVs or SNVs of unknown clinical significance, which were deemed not to be relevant to their neurodevelopmental phenotypes.

### Neurodevelopmental phenotypes

All the patients in the present cohort except for two (38/40, 95%) and all previously reported individuals had a diagnosis of developmental delay and/or IDD. The degree of impairment was variable (mild to severe/profound), with the majority having moderate IDD (Table 1). Comparing mutation classes and IDD severity, severe IDD was more frequent in PTVb class variants. Overall, there was a significant difference between groups (Kruskal-Wallis test, *p=0*.*0369*), however post hoc pairwise analysis (Mann-Whitney) between PTVa, PTVb and MISS groups (insufficient data for CNV and SPL) did not reach statistical significance, and the difference was not significant after including previous reported individuals in the combined cohort (p*=0*.*1632*, Supplementary Figure S2D).

Two affected individuals with PTVb class variants had been assessed as having normal cognitive functioning, with IQ 80 and 93; the first had dyslexia and dysgraphia and the second received speech therapy and special education. Both had a diagnosis of autism spectrum disorder (ASD).

Speech was affected in most individuals (Table 1), and often involved both receptive and expressive language. Median age at first words in the present cohort was 2 years (range 9m - 4^+6/12^y). Eight patients (and 2 previously reported) had absent speech at last evaluation. Dysarthria/articulation problems/speech apraxia were reported in 7 (and in 5 previously published). Gross and fine motor development were delayed in most individuals. Independent sitting was acquired at a median age of 11 months in the present cohort (range 6-19m). Median age at walking, either independent or with support, was 2^+4/12^ years (range 1^+2/12^-9y). There was no significant difference between variant classes with regards to age at first words or walking.

ASD was diagnosed in 35% of patients (34% in the combined cohort). There was a greater (non-significant) prevalence of ASD in patients with PTVb class variants in comparison with PTVa (44 vs 23% in the combined cohort; *p=0*.*2944*). Additional behavioral problems were reported in 67% of patients. They included: aggressiveness (towards self or others) in 36%, repetitive behavior in 33%, sleep disturbances in 30%, attention deficit hyperactivity disorder in 26%, and anxiety in 10%. Frequencies when including previously reported individuals were identical (Supplementary Table S1).

### Additional neurologic manifestations

Twenty-one percent of the patients in the present cohort had seizures (24% in the combined cohort). Two additional individuals had suffered a single seizure (P13 and P38). Median age at onset was 4^+3/12^ years (range 5 m - 10 y). Seizures were polymorphic and included generalized tonic-clonic, myoclonic, and focal-onset, with no dominant seizure type (Supplementary Table S1). Insufficient information was available regarding frequency and response to anti-epileptic drugs (Supplementary Table S1). Of note, 2 patients without clinical seizures had abnormal EEG findings.

A single individual with a PTVa class variant had seizures, vs. 35% in the PTVb class (difference not statistically significant, *p=0*.*2103*). In the combined cohort, more patients with PTVb class variants (31%) than PTVa class (20%) had seizures, though this difference was not significant. Half of all patients with missense mutations had seizures, though the total number of individuals reported was low (4 and 6 in the present and combined cohorts, respectively). None of the 3 patients with splice variants with information available had seizures.

Hypotonia was seen in 65% of the patients in the present cohort (70% when including previously reported patients). Hypotonia was more frequent in individuals with PTVa variants affecting all three isoforms when compared to MISS (*p=0*.*0410*) and to PTVb, though the latter was only significant when considering the combined cohort (*p=0*.*0495*).

Motor impairment (ataxia, broad based gait or lower limb spasticity) was reported in 45% of individuals of the present cohort (54% in the combined cohort).

### Hindbrain abnormalities are common in BCL11A-IDD

Brain abnormalities were identified on MRI in 58% of patients (22/38); 61% in the combined cohort (Supplementary Table S1). The most common findings were cerebellar abnormalities (34%; 28% in the combined cohort) and brainstem abnormalities (32%; 31% in the combined cohort) (Figure 3). A small or hypoplastic cerebellar vermis was observed in all of these (Supplementary Table S4). BCL11A/CTIP1 is highly expressed in Purkinje cells of the rodent cerebellum.^4^ Single cell gene expression analysis similarly showed high expression of BCL11A in Purkinje cells (PC) of the human cerebellum^30,31^, yet protein expression beyond Carnegie stage 19 in human development had not been explored. Here, we used immunohistochemistry to confirm that protein expression is high in the PC layer in human development from 12 to 18 pcw (Figure 4A-C). The primary brain stem imaging finding was a small pons objectively or relative to medulla, noting BLC11A is also expressed in the developing human brainstem (Aldinger et al. 2021,^30^ Figure 4D). Comparing the frequency of cerebellar and brainstem abnormalities with those identified in large contiguous gene deletions (Supplementary Table S5), there was no significant difference.

**Figure 2.**
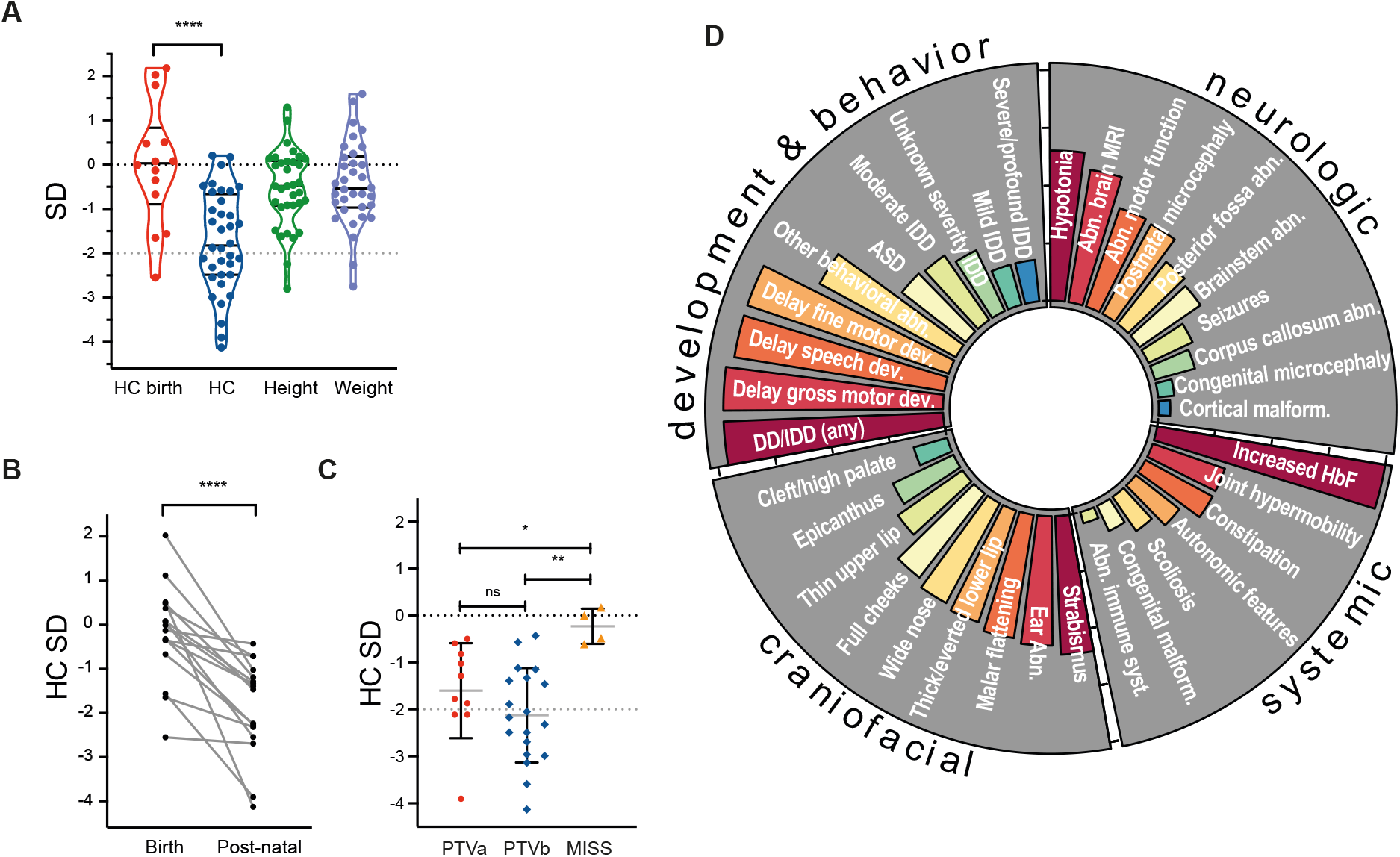
Clinical features of individuals with BCL11A-IDD in the present cohort. **A**. Distribution of growth parameters (standard deviations, SD) of present cohort (a single value for each individual is represented); lines indicate median, upper and lower quartiles; **** unpaired t-test p<0.0001. n=14 HC birth; n=36 HC; n=34 height & weight. **B**. Head circumference (HC) at birth and postnatal (SD for age and sex) for individuals in present cohort and previously reported where both measurements available; n=16 (13 PTV, 1 MISS, 2 CNV); **** 2-way ANOVA p<0.0001, F(1,15)=32.50. **C**. Head circumference SDs for PTV classes a (n=9) and b (n=19) and MISS (n=4). Unpaired t test: * p=0.0238; **p=0.0015; ns, not significant. **D**. Circular bar plot representing frequencies for phenotypic features in the present cohort; bars are proportional to frequency.

**Figure 3.**
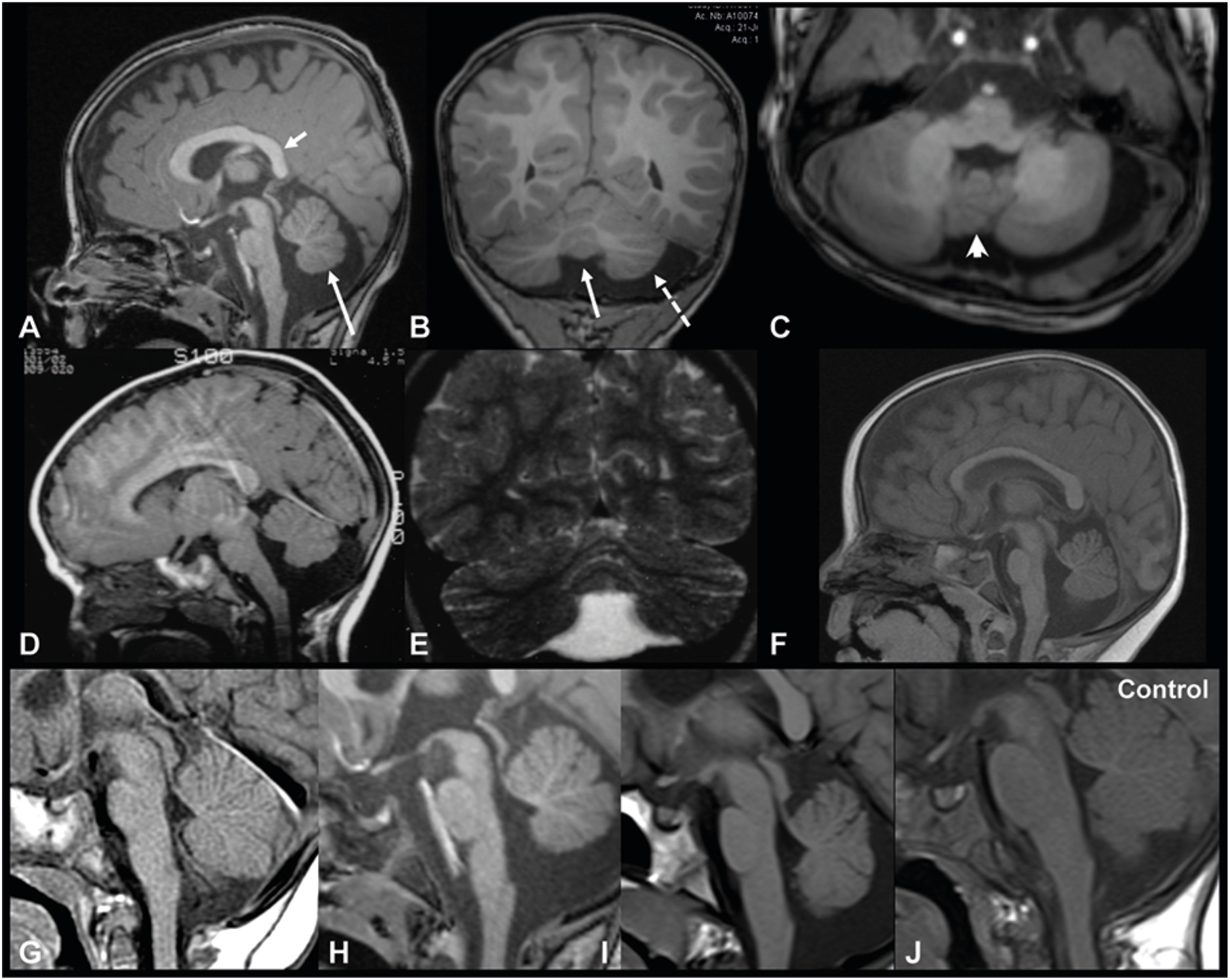
Neuroradiological features of individuals with BCL11A-IDD. Sagittal (**A**), Coronal (**B**) and axial (**C**) 3D T1 weighted-images (WI) in P34 showing inferior vermian hypoplasia (long arrows in A and B) with associated enlargement of the tegmento-vermian, mild reduction in size of the posterior aspect of the corpus callosum (short arrow in A), hypoplasia of the left cerebellar hemisphere (dashed arrow in B) and mild dysplasia of the vermis (arrowhead in C). Sagittal T1 WI (**D**) and coronal T2 WI (**E**) in P24 and sagittal T1 WI in P8 (**F**) shows isolated vermian hypoplasia with associated enlargement of the tegmento-vermian. Vermian hypoplasia was confirmed with measurements compared to normal values described in ref. ^28^. The pons is small in both the patients. Sagittal T1 WI in P15 (**G)**, P34 (**H**), and P41 (**I**) show the short pons and abnormally elongated medulla. Normal for comparison on the right (**J**).

**Figure 4.**
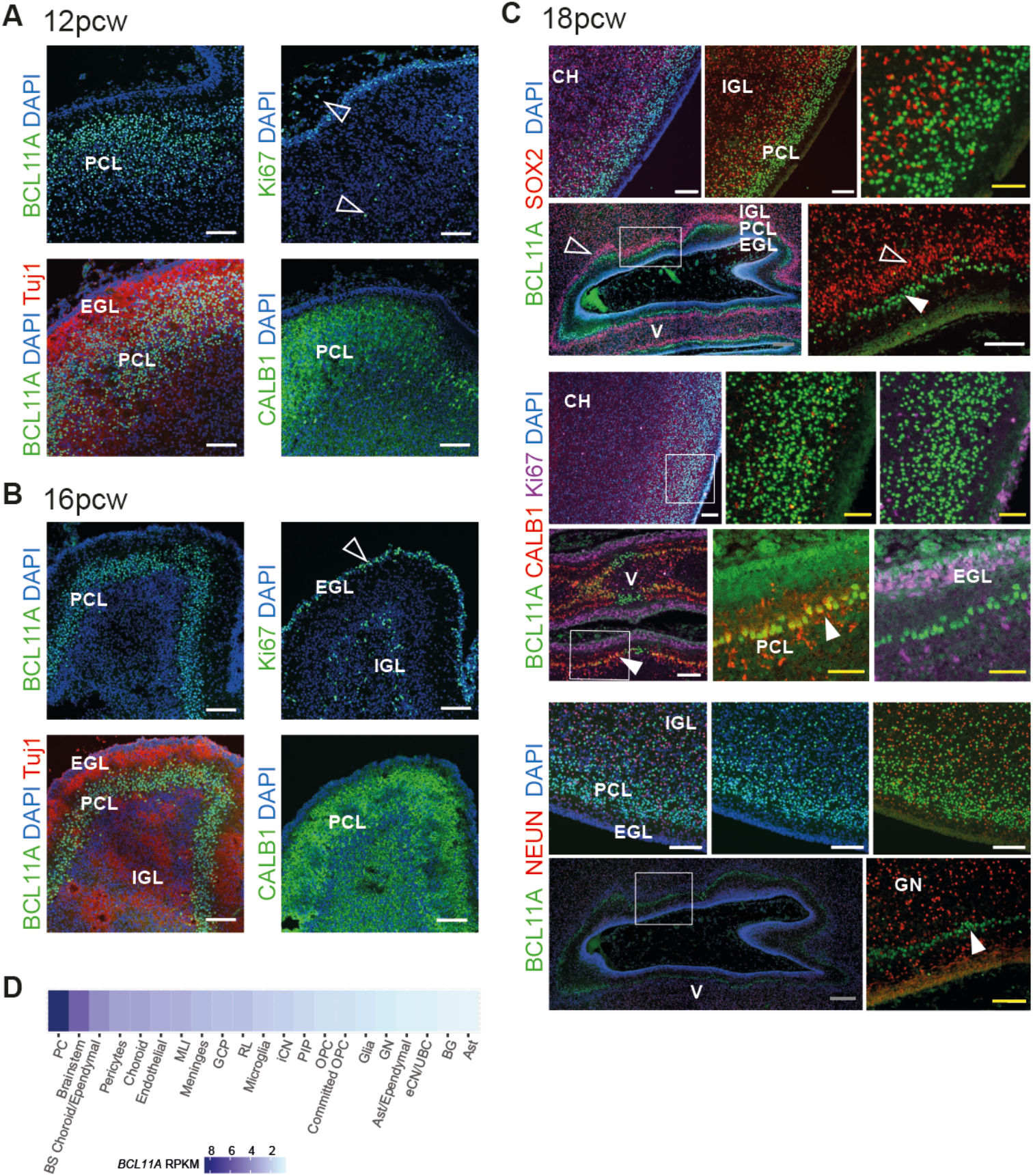
BCL11A expression in developing human cerebellum. Immunohistochemistry of human fetal cerebellum at 12 (**A**), 16 (**B**) and 18 (**C**) weeks post conception. CH, cerebellar hemisphere; V, developing vermis (with island like fissures); filled arrow, Purkinje cells; open arrow, cerebellar granule neuron precursors; EGL, external granule layer; PCL, Purkinje cell layer; IGL, internal granule layer; GN, granule neurons. Bars: grey, 200µm; white, 100µm; yellow, 50µm. **D**. BCL11A normalized gene expression by cell type in developing human cerebellum (data from ref. Aldinger et al., 2021^30^). RPKM, reads per kilobase of transcript per million mapped reads.

Cortical malformations were rare, occurring in only 2 patients in the present cohort: bilateral small frontal cortical dysplasia; left fronto-insular closed lips schizencephaly (Supplementary Table S4), with no additional cases in the previously reported individuals. For the former, images were not available for review. Compared to patients reported with large contiguous gene deletions (Supplementary Table S5), there were significantly less cortical malformations in both the present (*p=0*.*0260*) and the combined cohort of BCL11A-IDD (*p=0*.*0117*). Seven patients (18%) had callosal abnormalities, including hypoplastic or dysmorphic corpus callosum (19% in the combined cohort), a frequency identical to that observed in reported large CNVs (19%).

### Craniofacial phenotype

On the consensus phenotype recorded, shared subtle dysmorphism were noted, though a distinctively recognizable facial appearance was not identified (Table 1). We found that the most common facial features in patients from the present cohort were: abnormalities of the external ear (56%); malar flattening (54%); thick or everted vermillion of the lower lip (51%) and thin vermillion of the upper lip (34%); wide nose (49%); full cheeks (46%); epicanthus (29%) (Supplementary Table S1). Ear abnormalities were variable, most commonly small and/or attached earlobes. There was no significant difference between variant classes with respect to facial features, except for malar flattening, which was significantly more frequent in individuals with PTVa compared to patients with PTVb variants in the combined cohort (*p=0*.*0464*) (although not significant when restricted to the present cohort *(p=0*.*1194)*).

Signs of increased body hair (i.e. synophrys, thick eyebrows, long eyelashes, hypertrichosis) were recorded in 10/41 (24%) patients in the present cohort (12/53, 23% in the combined cohort), with no significant difference between variant classes. Frontal upsweep of hair was noted in 4 individuals from the present cohort and in one previously reported.^12^

Where information at different ages was available, we noticed that the facial appearance changed over time. While a round face with full cheeks and thick/everted vermillion of the lower lip were more evident at younger ages, the face tended to become longer and the facial dysmorphism less recognizable in adolescence and young adulthood.

Sixty percent of the affected individuals (in the present cohort and when including previously reported patients) had strabismus, with no significant difference among variant classes.

### Prenatal and birth history

Pregnancy was uneventful in most affected individuals in the present cohort (27/38, 71%). In 11 patients (29%) variable abnormalities in the prenatal history were reported (detailed in Supplementary Table S1). No problems at birth were reported in most patients, except for 2 individuals (and 1 previously reported), who presented with perinatal distress.

### Congenital malformations

Congenital malformations were seen in 5/41 patients (12%), including polydactyly, cleft palate, stenosis of the pulmonary artery branches, craniosynostosis and umbilical hernia. The frequency of congenital abnormalities in the present cohort is significantly lower than in reported individuals with large CNVs encompassing additional coding genes (Supplementary Table S5, 12/20; *p=0*.*0002*). This difference remained significant in the combined cohort (10/65; vs. large CNVs *p=0*.*0002*).

### Growth

In all patients where head circumference (HC) at birth was available, measurements were within the normal range, except for one individual with congenital microcephaly. Median HC SD was 0.03 (range -2.55 to 2.18 SD). Postnatal microcephaly (<-2SD or <3^rd^ centile) was observed in 16/36 (median of available values -1.83 SD, range -4.13 to 0.21; Table 1; Figure 2A). In the combined cohort, congenital microcephaly was observed in no additional patients and postnatal microcephaly was observed in 26/52 (median of available values -1.88 SD, range -4.42 to 0.21; Table 1; Supplementary Figure S2A).

Postnatal HC for age was significantly smaller than at birth (median SD 0.03 vs -1.83, *p<0*.*0001*) (Figure 2A). The same trend remained when including previously reported individuals (Supplementary Figure S2A). Analyzing individuals where HC measurement was available for both timepoints confirmed progressive relative postnatal microcephaly, with postnatal head size significantly smaller than at birth (*p<0*.*0001*, Figure 2B). Individuals with PTV had significantly smaller head size compared to those with MISS variants (*p=0*.*0207* and *p=0*.*0014* for PTVa and PTVb vs. MISS, respectively; Figure 2C), and those with PTVb class variant had overall smaller head size within the cohort (median SD -2.124, 95% CI [-2.609, - 1.640]).

Most patients had height and weight within normal ranges for age (Supplementary Table S1), with only 2 patients with short stature (SD <-2SD), and an additional 2 reported individuals; 3 reported individuals have short stature, but values are unavailable. Median height and weight SDs in the present cohort were -0.48 (95% CI [-0.87, 0.04]) and -0.54 (95% CI [-0.85, 0.00]), respectively. There were no significant differences for height and weight between variant classes (Supplementary Figure S2C), though on average patients with PTVb class variants were shorter.

### Hematological phenotype

Fetal hemoglobin (HbF) above maximum reference for age was observed in all individuals where hemoglobin electrophoresis or high-performance liquid chromatography (HPLC) was performed [median age at last HbF measurement: 10 year^+6/12^ (range 5-19)]. Median value of HbF was 15.9%, with a range of 4.1-35 (maximum reference value for individuals over 6 months of age: 2%). There was an inverse trend between age and HbF (Supplementary Figure S2E). In a subset of individuals where sequential measurements were available, HbF persistence decreased over time (Supplementary Table S6), yet always remaining above reference levels for age.

BCL11A is highly expressed also in B-lymphocytes, and BCL11A-deleted adult mice have impaired lymphoid development^32^. To assess the relevance of this to individuals affected with BCL11A-IDD, we evaluated the presence of immunological issues. Only one patient exhibited abnormalities of the immune system (P38, Supplementary Table S1). This individual was diagnosed with hypogammaglobulinemia, had recurrent pneumonias since the age of 6 years, and received treatment with intravenous immunoglobulins (Ig). In 4 other patients where an Ig panel was performed, this was normal.

### Additional features

Scoliosis was noted in 18% of affected individuals (23% in the combined cohort). When restricting the analysis to patients older than 10 years of age, scoliosis was seen in 4/10. Joint hypermobility/laxity was reported in 33% of patients (38% in the combined cohort). Signs of autonomic dysfunction were reported in 24% of patients. They consisted mostly of cutis marmorata or telangiectasia, intermittently cold extremities or Raynaud’s phenomenon, and reduced/increased sensitivity to pain. Thirty-three percent of patients had constipation (38% in the combined cohort). There was no significant difference between variant classes with regards to these manifestations.

Because somatic variants in *BCL11A* have been reported in single sporadic malignant tumors^33^ and altered *BCL11A* expression has been detected in lymphomas^34^, we queried the presence of malignancies in patients with BCL11A-IDD. None of the affected individuals had been diagnosed with either solid or hematologic malignancies. Only one patient had a benign tumor (osteochondroma).

## Discussion

We present the detailed phenotype of 42 patients with BCL11A-IDD and expand on the phenotypic spectrum by reviewing an additional 33 previously reported individuals. Our study provides the first comprehensive delineation of the phenotypic and genotypic spectrum of BCL11A*-*IDD thus far based on information of these 75 affected individuals. We find some evidence for correlation between genotype and severity of phenotype. Though not all parameters measured reach statistical significance, we see a trend towards a more severe phenotype of protein truncating variants (PTV) in comparison with missense and splice variants. The previous evidence for a hypomorphic effect of missense variants (3 previously investigated functionally)^5^ and the persistence of fetal hemoglobin in all patients with MISS variants tested are consistent with a loss of transcriptional repression activity. We further dissected the genotypic analysis by comparing variants predicted to affect all three primary isoforms (PTVa) and those sparing BCL11A-S (PTVb). Unexpectedly, we found that for PTVb there is a trend towards more severe neurologic features, namely severity of developmental delay, ASD, and seizures, when comparing with either PTVa or other variant types. Microcephaly was significantly more severe in PTVs compared with MISS variants, more so for PTVb. Only hypotonia and discrete facial features were significantly different in PTVa mutations.

The identification of two patients who inherited their variants from an apparently unaffected parent suggests intrafamilial variability or possibly reduced penetrance, although HbF was not measured in the parents. One of them (P29) is one of two individuals with cognitive abilities in normal range; however, both cognitively normal individuals had speech and language difficulties. Ratio of the mutant and wildtype allele in blood of P29 and parent make the possibility of mosaicism in the parent unlikely. It is also possible that adult individuals with mild IDD or history of speech impairment may not be readily diagnosed unless formally assessed. These cases highlight the importance of testing parental samples to ensure appropriate recurrence risks and genetic counselling as well as detailed assessment of parental phenotypes in such cases. We also provide the first two reports of apparent germline mosaicism in BCL11A-IDD, where the parents did not carry the variant in blood or saliva yet had recurrence of the disease in their offspring.

Disregarding the possibility of NMD escape and assuming equal allelic expression, heterozygous PTVb variants would result in a disproportion of BCL11A-S isoform in relation to BCL11A-L and -XL. Our and others’ previous work has shown that wild-type BCL11A-L and BCL11A-S form homo- and heterodimers^5,35^, and that co-expression of BCL11A-S and -L isoforms lead to translocation of BCL11A-S from the cytoplasm into nuclear paraspeckles^5^. We speculate that -L (and -XL) -specific haploinsufficiency leads to decreased availability of protein for heterodimerization with -S, thus decreasing its putative nuclear function. Furthermore, BCL11A-S is thought to negatively regulate BCL11A-L by preventing its homodimerization^36^, inhibiting its negative control of axon branching and dendrite outgrowth, and overexpression of BCL11A-S has a cytotoxic effect on cultured rat hippocampal neurons. This suggests that PTVa variants that spare BCL11A-S and cause haploinsufficiency of -L and -XL isoforms could have a dominant-negative effect.

Conversely, all but 5 variants may be subject to NMD escape (Supplementary Table S2). Given the structure of the three BCL11A isoforms, they are likely to be subject to NMD by different mechanisms, and in some cases more than one escape mechanism may be at play^27^. That would result in significantly truncated proteins, with (frameshift) or without (stop) aberrant protein sequence. Considering that possibility, we compared phenotypic frequencies between stop and frameshift mutations and did not identify differences. The possibility remains that truncated BCL11A-L and -XL isoforms are translated, evade proteasome degradation, and have a gain of function of toxic effect. If that were the case, we would expect a progressive phenotype. To our knowledge, there have not been any reports of regression or neurodegeneration to date, though we appreciate long term follow-up of adult individuals would be required to exclude this.

The latter possibility also seems less likely considering recent evidence for altered NMD mechanisms in rare variants. Using allele-specific expression, Teran et al. (2021) created predictive models of NMD efficiency, finding that rare and ultra-rare variants (MAF <0.001%) are less likely to escape NMD.^37^ Variants classified as pathogenic in ClinVar were more likely to exhibit allelic imbalance due to NMD. Allelic imbalance affecting rare variants was determined by variant location, allele frequency, and conservation, remaining consistent across tissue types. Further investigations will be required to test these hypotheses for patient-specific BCL11A variants.

Interestingly, in the <u>gnomAD</u> database (v.2.1.1)^38^, which excludes known severe pediatric disease, there are only 8 variants potentially truncating at least one isoform recorded (Supplementary Table S7). All but one (p.Ser248Ter) are in the C-terminal region of the protein, downstream of the variants identified in the BCL11A-IDD cohort, and downstream of a putative TBR1 interaction domain (residues 629–773 of BCL11A-L; Supplementary Figure S1A)^39^, an interacting transcription factor crucial for neurodevelopment. Given the variable severity of IDD and paucity of congenital malformations in BCL11A-IDD identified in our cohort, we posit that either a) the individuals may have been recruited as unaffected because of a mild neurodevelopmental phenotype, or b) the gnomAD variants have minimal phenotypic impact due to their C-terminal location.

Hindbrain abnormalities in BCL11A-IDD appear to be common, affecting at least a third of patients (Figure 2D). The neuroradiological findings can be subtle, as was the case for 2 patients reanalyzed, where previous clinical reports indicated a normal cerebellum. The hindbrain phenotype of a relatively small pons may be under-reported, as measurements are not routinely performed. The collection of findings in a subset of patients may represent a mild pontocerebellar hypoplasia phenotype, though should be distinguished from classical forms of pontocerebellar hypoplasia (PCH) by absence of evolving cerebellar atrophy and associated supratentorial features^40^. Interestingly, individuals with large CNVs encompassing additional coding genes do not have an increased frequency of posterior fossa abnormalities, even though overall they present with more congenital anomalies and cortical malformations. These observations together with the expression pattern of BCL11A in development suggest that: a) normal BCL11A expression is important, albeit not required, for the development of anatomically normal hindbrain, and b) BCL11A haploinsufficiency (and not adjacent coding or non-coding regions) is responsible for cerebellar and pontine anomalies in individuals with deletions including 2p16. Together with observations by Aldinger et al. (2019)^23^, our data supports the role of *BCL11A* as a hindbrain abnormalities gene.

On the other hand, given that cortical malformations are rare in BCL11A-IDD, this could suggest that other genes in the 2p15p16 region have a more significant role, or that neocortex development is less sensitive to BCL11A haploinsufficiency. In mouse models, BCL11A is required for late differentiation and survival of upper layer neurons, and conditional biallelic loss (constitutively lethal) is associated with significant migration defects of upper-layer projection neurons and abnormal deep layer neuron specification^8^. Moreover, as referenced above, BCL11A N-terminus directly interacts with TBR1, a transcription factor critical for early born neurons in the developing cortex, in *in vitro* assays^39^. It is possible that heterozygous loss of *BCL11A* is not sufficient to cause an overt cortical malformation phenotype that can be detected by current imaging techniques, and as biallelic loss of BCL11A is not expected to be viable in humans, such phenotype would not be observed. Even so, half of patients have postnatal microcephaly, supporting a role for BCL11A in human cortical development. Its role is also supported by transcriptional deregulation in cortex (and hippocampus) of the heterozygous mouse model^5^. Of note, the presence of schizencephaly in one patient may be incidental and related to a prenatal ischemic damage, the most likely cause of schizencephaly.

In keeping with this, postnatal microcephaly is a common and previously underappreciated feature of BCL11A-IDD, present in half of the affected individuals and with HC as low as -4 SD. Even in normocephalic patients, HC is below average for the general population in most (Supplementary Table S1, Supplementary Figure S2A).

BCL11A has been reported as a putative subunit of the BRG1- and BRM-associated factor (BAF) swi/snf chromatin remodeling complex ^33^, having been identified by mass spectrometric analysis in highly purified BAF complexes in human T cells and P1.5 mouse brain. The BAF swi/snf complex plays multiple roles in brain development^41^, most notably regulating the transition between neural progenitors to post-mitotic neurons^42,43^. Mutations in BAF swi/snf subunits are an important cause of neurodevelopmental disorders, including Coffin-Siris (CSS, e.g., mutations in *ARID1B, SMARCB1, ARID1A, SMARCA4, SMARCE1, ARID2*) and Nicolaides-Baraitser syndromes (NCBRS, *SMARCA2* mutations)^44,45^. In addition to IDD, patients share physical features such as coarseness, integumentary abnormalities, including sparse hair and body hirsutism, and digital anomalies. More recently, expanding phenotypes (e.g. *SMARCA2, SMARCA4*) ^46,47^ and novel BAF swi/snf genes involved in syndromic and non-syndromic IDD (e.g. *ACTL6A, ACTL6B, SMARCC1*)^48-51^ without classical CSS and NCBRS physical features have broadened the spectrum of conditions considerably. The phenotypic variability in swi/snf related disorders may be related to the different combinatorial assemblies of BAF, with developmental stage and tissue specific incorporation of different subunits, or to the different DNA binding and epigenetic mark recognition properties of the vast repertoire of BAF proteins^52^. Interestingly, subsequent to the study by Kadoch et al. ^33^, to our knowledge no further evidence supporting BCL11A as a bona fide BAF swi/snf subunit has been reported. Further investigations are required to confirm the interaction between BCL11A and BAF, and its relevance to brain development. We speculate that rather than a subunit, BCL11A may be a BAF transcription factor interactor in a context specific manner. BAF swi/snf complexes interact with and/or are recruited by sequence specific transcription factors that direct lineage-specification and differentiation^52^. A similar occurrence is that of ADNP, a homeodomain-containing zinc finger protein that interacts with BAF swi/snf complexes, and is also associated with IDD and ASD through a haploinsufficiency mechanism^53^.

Over a third of affected individuals are diagnosed with ASD, and additional behavioral problems are seen in more than two thirds of patients (Figure 2D). Although children with BCL11A-IDD are usually described as friendly, burst of aggression were reported in 35%, which can be related to underlying behavioral pathology or expressions of frustration due to difficulties in communicating. Therefore, we recommend formal behavioral and ASD assessments (e.g. ADOS) for all patients with BCL11A-IDD to ensure appropriate support. While dysarthria and/or articulation problems were reported in 7 patients from the present cohort, further studies are needed to confirm the suggestion that *BCL11A* is a candidate gene for childhood apraxia of speech^54^.

Seizures have been reported occasionally in individuals with BCL11A-IDD^14,16^, but the prevalence had not been investigated. Here we show that seizures occur in 25% of patients, usually starting in early childhood. There does not seem to be a recurrent seizure type or treatment-response pattern, though more detailed assessment of larger cohorts and functional research are needed to determine whether disease-specific therapeutic regimens can be proposed.

Hypotonia was seen in most affected individuals. Interestingly, ataxia and/or spasticity were reported in 45% of individuals from the present cohort and in 54% in the combined cohort. This number may be affected by a selection bias, as we note the inclusion of 4 patients listed in ClinVar as having IDD and spastic paraplegia that were submitted by the same center (Supplementary Table S1). Nonetheless, it is possible that a subset of patients who present with hypotonia develop motor impairment and spasticity with age.

As seen in several neurodevelopmental disorders, BCL11A-IDD is not associated with a recognizable facial appearance. Nonetheless, recurrent subtle dysmorphism were reported, and facial features such as malar flattening, full cheeks and full or everted lower lip are commonly seen in affected individuals. Strabismus is seen in 2/3 of the patients, warranting at least a baseline ophthalmologic evaluation in all affected individuals.

Congenital malformations in individuals with single nucleotide variants or CNVs encompassing only *BCL11A* were rare. The finding of polydactyly in 4 individuals (postaxial in at least 2) was unexpected (Supplementary Table S1). BCL11A is expressed in mouse developing limb buds^5^, however limb anomalies have not been observed in the mutant mouse models^5,32^. It is unclear whether these are a direct effect of *BCL11A* haploinsufficiency. Experimentation in animal models may be needed to define the role of *BCL11A* in developing limbs and ensuing pathology.

Conversely, overall birth defects were significantly more frequent in patients with large CNVs encompassing additional coding genes, suggesting that other genes in the deleted region may be responsible for congenital malformations. This finding strengthens the hypothesis that BCL11A-IDD and larger deletions of 2p15p16.1 encompassing *BCL11A* with multiple adjacent genes should be considered two distinct conditions^1^, although *BCL11A* haploinsufficiency certainly plays a crucial role in the 2p15p16.1 deletion syndrome phenotype.

Features suggestive of autonomic dysfunction are newly detected manifestations of BCL11A-IDD. Interestingly, in 1981 Manders et al. reported on a child with IDD, persistent HbF and Raynaud’s phenomenon^55^, which could represent the very first description of BCL11A-IDD or 2p15p16.1 deletion syndrome. Autonomic dysregulation has been described in other neurodevelopmental disorders such as Rett and Pitt-Hopkins syndromes^56,57^. Therefore, we suggest clinicians proactively query changes in extremities appearance, gastrointestinal dysmotility and possible other signs of autonomic dysregulation in individuals with BCL11A-IDD.

We and others had previously reported on elevated HbF in association with BCL11A variants and microdeletions^5,11,58^. BCL11A is a transcriptional repressor of HbF that acts by repressing the *HBG1/2* genes encoding γ-globin (the fetal β-like globin) and plays a critical role in globin switching from fetal to adult hemoglobin (HbA) as part of a complex regulatory network^59^. Like hereditary persistence of fetal hemoglobin (HPFH), a benign condition usually caused by deletions encompassing the β-globin gene cluster or by point mutations in the γ-globin gene-promoter region, it is thought that elevated HbF itself is a benign feature in BCL11A-IDD. In the present cohort no other hematological abnormalities were identified on routine investigations, though prospective assays were not performed (apart from 2 patients). In individuals affected by β-thalassemia and sickle cell anemia, elevated HbF may mitigate symptoms, and thus BCL11A-silencing is being actively pursued as a potential treatment for hemoglobinopathies^60^. However, HPFH should be easily distinguished from BCL11A-IDD as it lacks the cognitive and systemic features discussed above. As previously identified in individuals with HPFH in association with sickle cell anemia, we identified a negative correlation between HbF levels and age^61^, though the significance of this finding in the absence of concomitant hematological disease is unknown.

As increased HbF is a hallmark of BCL11A-IDD, we advocate using HbF testing by hemoglobin electrophoresis or HPLC as a diagnostic tool, particularly for variants of unknown significance. Attention should be given to age because of the physiologically elevated HbF in the general population in the first 12-24 months of life^62^, which precludes it as an early infant biomarker, namely in conjunction with newborn screening. We also recommend that for individuals incidentally diagnosed with persistence of HbF that have neurodevelopmental concerns, *BCL11A* mutation analysis should be performed. Most individuals will have undergone chromosomal microarray, exome or genome sequencing for IDD diagnostic investigations, thus ruling out β-globin gene cluster deletions and common γ-globin polymorphisms associated with HPFH. On the other hand, BCL11A-IDD can mask HPFH, whereby elevated HbF has dual etiology. Hence, parental HbF should be offered for family counselling regarding possible thalassemia risk.

In this large cohort, neither signs of immunological dysfunction nor increased risk for malignancy were observed. Despite this encouraging data, we note that most patients in this series are young, making it impossible to detect age-dependent phenotypes. A prospective analysis of these patients throughout adulthood or identification of larger numbers of affected adults will be required to determine additional features, e.g. progressive neurologic manifestations or scoliosis, and to exclude putative malignancy risk. We recognize also that the inclusion of patients from publicly available repositories where limited information is available may over or underestimate the frequency of some features.

In summary, we provide a comprehensive delineation of the BCL11A-IDD phenotype in the largest cohort reported thus far. We expand the phenotypic spectrum to include previously underappreciated manifestations such as postnatal microcephaly, behavioral abnormalities, seizures, abnormal motor function, and signs of autonomic dysregulation. A detailed analysis of brain imaging detected cerebellar and posterior fossa abnormalities as a prevalent manifestation BCL11A-IDD, supporting the role of *BCL11A* in hindbrain development. Our study also expands the *BCL11A* mutational spectrum and indicates that protein-truncating variants tend to be associated with a more severe phenotype compared to the other mutation types, and that BCL11A-S retention may contribute to protein dysfunction. Additional investigation into the pathogenesis of disease in human *in vitro* models will be required to further characterize the molecular and cellular defects of BCL11A in human brain development.

## Data Availability

Clinical and molecular data included in this study will be made publicly available at www.bcl11agene.com on peer-reviewed publication.
Genetic analyses: for access to the retrospectively performed analyses, please contact the authors who will refer to the appropriate center (data availability is subject to each individual study's ethics and consent).

## Data availability

All clinical and molecular data included in this study are provided in Supplementary Table S1 and will be made publicly available at www.bcl11agene.com/ on publication.

Genetic analyses had been previously performed. For access, please contact the authors who will refer to the appropriate center (data availability is subject to each individual study’s ethics and consent).

## Online resources

www.ensembl.org/

www.bcl11agene.com/

https://deciphergenomics.org/

https://omim.org/entry/617101

https://gnomad.broadinstitute.org/gene/ENSG00000119866?dataset=gnomad_r2_1

## Acknowledgments

We would like to acknowledge the patients and their families. We would also like to acknowledge the Deciphering Developmental Disorders (DDD) Study. The DDD study presents independent research commissioned by the Health Innovation Challenge Fund [grant number HICF-1009-003]. This study makes use of DECIPHER (http://decipher.sanger.ac.uk), which is funded by Wellcome. See Nature PMID: 25533962 or www.ddduk.org/access.html for full acknowledgement. We thank Daniel Perrett of the DECIPHER team for assistance in generating Figure 1 and Supplementary Figure S1A. Part of the data presented here was provided through access to the data and findings generated by the 100,000 Genomes Project, which is funded by the NIHR and NHS England (full acknowledgement on https://www.genomicsengland.co.uk/about-gecip/publications/). We acknowledge the contributions of the C4RCD Research Group (members are listed in supplementary data). The Center for Rare Childhood Disorders is funded by private donations made to the TGen Foundation. We acknowledge the contributions of the TUDP consortium (Telethon Undiagnosed Disease Program; members are listed in supplementary data). We thank the Francis Crick Institute Experimental Histopathology team and Clementina Cobolli-Gigli for experimental support, and Simona Amenta and Odera Aguele for clinical support.

C.D. is supported by the Wellcome Trust [grant number 209568/Z/17/Z]. The BUILD Study is supported by the Wellcome Trust [grant number 209568/Z/17/Z], the NIHR UK Rare Genetic Disease Research Consortium and The Great Ormond Street Hospital NIHR Clinical Research Facility. A-L.B. work was supported by grants from the Regional Council of Burgundy (Plan d’Actions Régional pour l’Innovation—PARI) and the fonds européen de développement regional (FEDER). B.B.A.d.V’s work was financially supported by grants from the Dutch Organization for Health Research and Development (ZON-MW grants 917–86–319 and 912–12–109). D.B’s work was supported by the National Cancer Institute of the National Institutes of Health under Award Number R01CA210561, Akhurst PI (DTB). M.J.B. and the UW-CMG are supported by NHGRI and NHLBI grants UM1 HG006493 and U24 HG008956, by the Office of the Director, NIH under Award Number S10OD021553. The content is solely the responsibility of the authors and does not necessarily represent the official views of the National Institutes of Health.

The authors acknowledge providers of human fetal material: Joint MRC/Wellcome Trust (grant# MR/R006237/1) Human Developmental Biology Resource (www.hdbr.org) and BDRL, University of Washington (NIH R24 HD000836). Human tissue used in this study was covered by material transfer agreements between The Francis Crick Institute and HDBR, and between Seattle Children’s Research Institute and HDBR/BDRL.

## Author Information

Conceptualization: A.P., C.D.; Data curation: A.P., C.D.; Formal analysis: A.P., C.D., F.D’A.; Investigation: A.P., C.D., F.D’A., P.H., M.J.G, W.B.D., D.V.; Funding acquisition: C.D., F.G.; Patient recruitment, clinical and diagnostic evaluations: A.P., F.D’A., K.A.A., C.S-H., C.Z., G.A.G., K.B., A.A., E.F.A., P.Y.B.A., R.B., D.B., L.M.B., A.B., A-L.B., Ø.L.B., P.M.C., V.C., C.C., J.C., A.C., J.C-S., D.L.E., L.F., C.P., P.F., L.G-N., D.H., S.H., G.H., J.H., C.K., B.E.K., J.L., K.J.L., S.A.L., S.M., R.M., R.M., C.M., K.McD., T.M., M.M., C.M., A.S.O., J.C.P., A.S.P., R.P., M.D., J.P., R.P., M.S., M.S-G., A.S., M.S., C.T., T.T., K.W., I.M.W., M.Z., C4RCD Research Group, TUDP, UW-CMG, B.B.A.deV., W.B.D., D.V., C.D.; Supervision: C.D.; Writing – original draft: A.P., C.D., F.D’A.; Writing – review and editing: all authors.

## Ethics Declaration

Ethical approval was obtained from the ethics committee of the following institutions: Deciphering Developmental Disorders Study: Cambridge South REC reference 10/H0305/83, and Republic of Ireland REC GEN/284/12; The 100,000 Genomes Project: Cambridge South REC 14/EE/1112; The BUILD Study: London - Camden & Kings Cross REC 17/LO/0981; Seattle Children’s Hospital IRB study #13291; Translational Genomics Research Institute, WIRB Protocol #20120789; Ethical review board of Friedrich-Alexander-University Erlangen-Nürnberg, #253_15B; University of California San Diego IRB protocol #101103; Rambam Medical Center IRB #0038-14-RBM; Ethics committee of Federico II University Hospital, protocol number 48/16; Johns Hopkins Medicine IRB protocol IRB00214093; Commissie Mensgebonden Onderzoek Regio Arnhem-Nijmegen NL36191.091.11. For clinically ascertained individuals IRB review was waived (by: Alberta Children’s Hospital REB; Comissão de Ética Hospital Pediátrico Centro Hospitalar e Universitário de Coimbra) and informed patient/guardian consent was given for publication of de-identified data and the appropriate institutional forms have been archived. For fetal samples, The HDBR ethical approval was granted by North East - Newcastle & North Tyneside 1 REC 18/NE/0290.

