## Supplementary for "BCL11A intellectual developmental disorder: defining the clinical spectrum and genotype-phenotype correlations"

Peron A. et al, 2021

This file includes:

A. Supplementary Figures S1 to S2.

B. Supplementary Tables S4, S6 and S8 (Supplementary Tables S1 to S3, S5 and S7 provided as separate files).

C. Supplementary Materials and Methods.

D. Supplementary Information – consortia authors.

E. Reference List

### A. Supplementary Figures S1 to S2

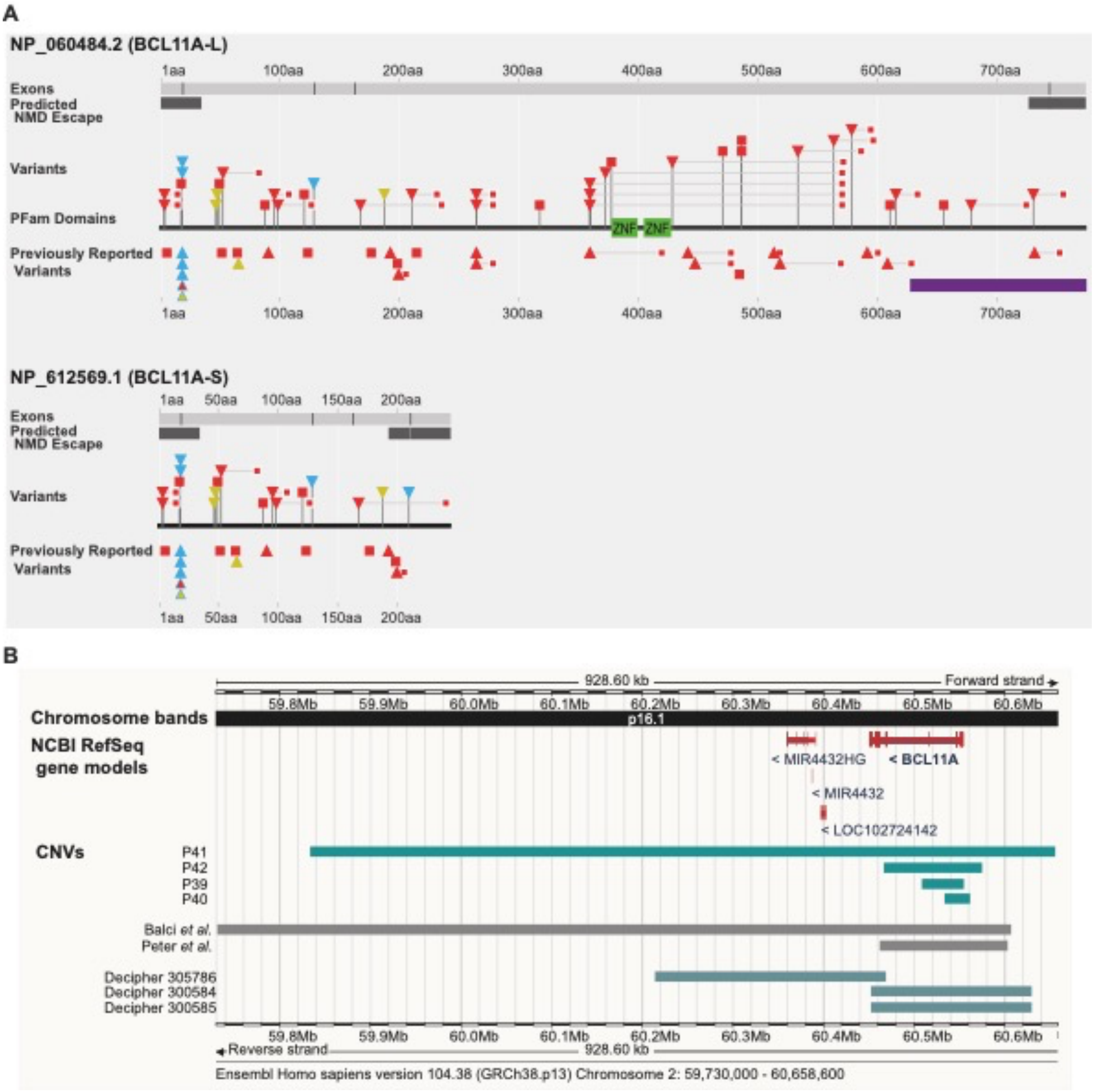

***Supplementary Figure S1. BCL11A*-IDD variants**; supplementary figure to Figure 1. **A.** Genomic variants annotated to the BCL11A-L (top) and BCL11A-S (bottom) isoforms. Stop gain: red square; frameshift: red triangle, with downstream premature termination codon as small red square; missense variants: yellow triangle; splice variants: light blue triangle. Light blue outline on red triangle: frameshift and potential splice variant ClinVar_VCV000987092.1; light blue outline on yellow triangle: missense and potential splice variant ClinVar_VCV000987093.1 (see Supplementary Table S2). Purple bar: region of putative interaction with TBR1[1]. Dark grey bars: predicted NMD escape by "start proximal" and "last exon junction". This figure was generated with the help of the Decipher team (https://deciphergenomics.org). **B.** CNVs presented in this study. Figure adapted from www.ensembl.org/.

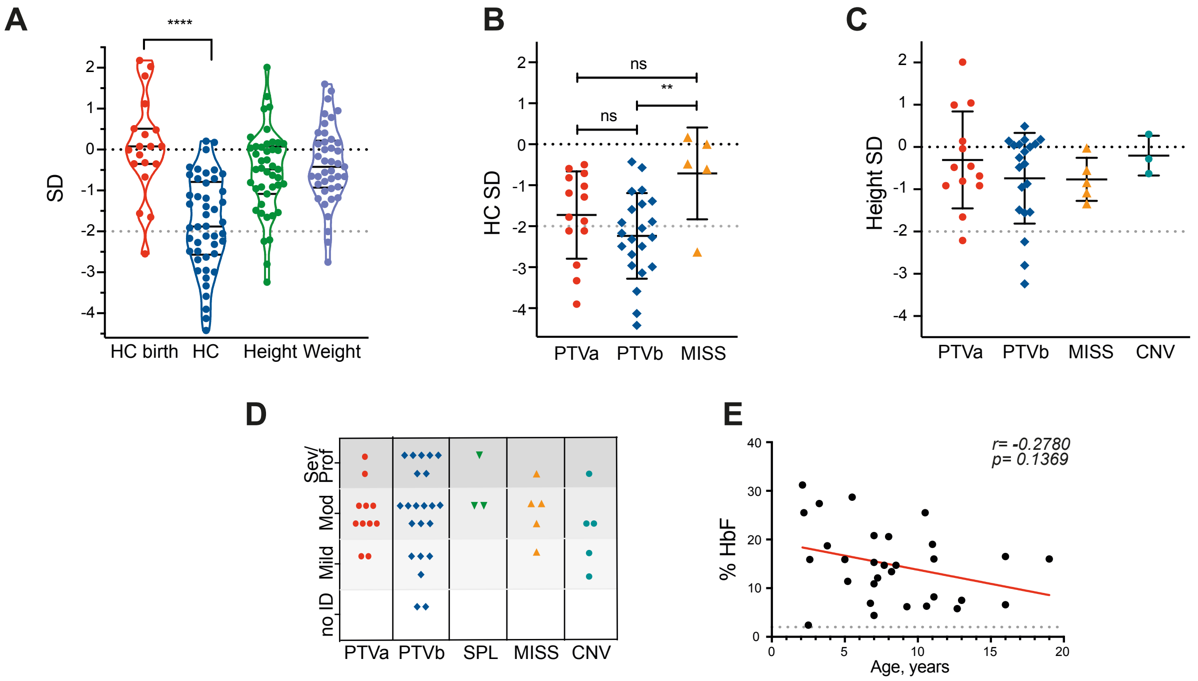

***Supplementary Figure S2.***

**Clinical features of individuals with BCL11A-IDD in the combined cohort.**

**A.** Distribution of growth parameters of present cohort and previously reported cases (a single most recent value for each individual is represented); lines indicate median, upper and lower quartiles; **** unpaired t-test *p<0.0001*. **B.** Head circumference SDs for PTV classes a and b and MISS; n=14 PTVa, 22 PTVb, 5 MISS; unpaired t test: ***p=0.0074*; ns, not significant. **C.** Height SD per mutation type; n=12 PTVa, 20 PTVb, 5 MISS, 3 CNV; 1 SPL not represented (data in Supplementary Table 1). **D.** Severity of intellectual disability per mutation type for patients in whom severity was available; for each degree of severity, the top row of datapoints represents the present cohort, the bottom previously reported cases; no significant difference between severity and mutation type (Kruskal-Wallis test, *p=0.1632* [for present cohort only, *p=0.0369*]; post-Hoc Mann Whitney tests not significant for p<0.05). **E.** Fetal hemoglobin (HbF) for age; all available data is presented (present and previously reported cohort; multiple measurements per individual where available); grey line: reference cutoff of 2%; red line: linear regression, $r^2=0.1048$; *p=0.*0810; Spearman correlation coefficient *r= -0.2780, p= 0.2473*.

### B. Supplementary Tables

**Provided as separate files:**

***Supplementary Table S1.*** Detailed clinical data of individuals reported.

***Supplementary Table S2.*** Sequence variants in *BCL11A* included in the full clinical cohort.

***Supplementary Table S3.*** Copy number variants in *BCL11A* included in the full clinical cohort.

***Supplementary Table S5.*** Summary of congenital malformations and brain MRI findings in previously reported individuals with large deletions encompassing additional genes[2-16].

***Supplementary Table S7.*** Loss of function sequence variants in *BCL11A* in GnomAD.

| Patient ID | Mutation, NP_075044.2: | Mutation class | Age range | Posterior fossa findings | Supratentorial findings |
|---|---|---|---|---|---|
| **P2** | p.Gly7AlafsTer9 | PTVa | 6-8y | small pons relative to medulla and in comparison with normal controls, small vermis height, mildly hypoplastic left cerebellar hemisphere | normal |
| **P5** | p.M53Pfs*32 | PTVa | ≤2y | vermian hypoplasia and small pons | small corpus callosum |
| **P8** | p.Val99TrpfsTer29 | PTVa | ≤2y | small pons relative to medulla and in comparison with normal controls, small vermis height | normal |
| **P11** | p.Gly212ArgfsTer21 | PTVb | 9-11y | small pons relative to medulla and in comparison with normal controls, small vermis height | dysmorphic corpus callosum |
| **P15** | p.Leu360ProfsTer21 2 | PTVb | 3-5y | small pons relative to medulla and in comparison with normal controls, small vermis height | dysmorphic corpus callosum, incomplete hippocampal rotation bilaterally |
| **P18** | p.Val374GlyfsTer198 | PTVb | NA[1] | normal | normal |
| **P24** | p.Val534AlafsTer54 | PTVb | NA[1] | small pons relative to medulla and in comparison with normal controls, small vermis height | normal |
| **P30** | p.Ser679GlnfsTer47 | PTVb | ≤2y | normal | minor white matter abnormalities |
| **P34** | c.385+2T>C | SPL | ≤2y | small pons relative to medulla and in comparison with normal controls, small vermis height, hypoplastic left cerebellar hemisphere, abnormal vermis foliation | dysmorphic corpus callosum |
| **P35** | p.Thr47Pro | MISS | ≤2y | normal | slightly reduced white matter volume |
| **P36** | p.Cys48Phe | MISS | 3-5y | small pons relative to medulla and in comparison with normal controls, small vermis height | normal |
| **P40** | partial gene deletion | CNV | ≤2y | slightly small pons on measurements | left fronto insular closed lip schizencephaly |
| **P41** | whole gene deletion | CNV | ≤2y | small pons relative to medulla and in comparison with normal controls, small vermis height, retrocerebellar cyst | dysmorphic corpus callosum |
| **Previously reported** | | | | | |
| **Dias_4** | p.Gln177Ter | PTVa | ≤2y | slightly small pons on measurements | normal |
| **Balci_1** | whole gene deletion | CNV | ≤2y | small pons relative to medulla and in comparison with normal controls, small vermis height and black's pouch cyst (BPC) | hydrocephalus associated to the BPC, bilateral malrotation of the hippocampi |

***Supplementary Table S4.***

Summary of brain MRI quantitative and qualitative analysis findings.

[1]Age at scan and full DICOM file not available; selected images analyzed only.

| Sex | Age range* (yrs) | HbF % | HbA1 % | HbA2 % |
|---|---|---|---|---|
| M | ≤2 | 25.5 H | 72.2 L | 2.3 |
| | ≤2 | 15.9 H | 82.0 L | 2.1 |
| | 3-5 | 11.4 H | 86.2 L | 2.4 |
| | 6-8 | 6.9 H | 90.6 L | 2.5 |
| | 9-11 | 6.2 H | 91.2 L | 2.6 |
| | 9-11 | 6.3 H | 91.2 L | 2.6 |
| F | ≤2 | 31.2 H | 67.4 L | 1.4 |
| | 6-8 | 20.6 H | 77.6 L | 1.8 |
| | 6-8 | 13.4 H | 84.3 L | 2.3 |
| | 6-8 | 14.7 H | 83.1 L | 2.2 |
| M | 9-11 | 8.2 H | n.a. | 2.5 |
| | 15-17 | 6.6 H | 79 L | 2.9 |
| F | 3-5 | 27.4 H | 71.4 L | 1.2 |
| | 3-5 | 28.7 H | 69.6 L | 1.7 |
| | 9-11 | 25.5 H | 72.8 L | 1.7 |
| **Previously reported** | | | | |
| F | 3-5 | 18.7 H | n.a. | n.a. |
| | 6-8 | 12.1 H | n.a. | n.a. |

***Supplementary Table S6.*** Sequential HbF measurements for a subset of individuals. H: high for age reference; L: low; n.a.: not available.

| Target | Host Species | Conjugate | Dilution | Company, catalogue number (clone ID), [RRID] | Tissue |
|---|---|---|---|---|---|
| BCL11A (Ctip1) | mouse | NA | 1:50 | Abcam, ab19487 (14B5), [AB_444947] | a), d) |
| BCL11A (Ctip1) | rabbit | NA | 1:100 | Abcam, ab19489 (18B12DE6), [AB_2063996] | b), c) |
| Calbindin-D-28-K | rabbit | NA | 1:100 | Sigma-Aldrich, C9848 (CB-955), [AB_476894 | d) |
| Calbindin D-28k | rabbit | NA | 1:3000 | Swant, CD38, [AB_10000340] | b), c) |
| Ki67 | mouse | NA | 1:50 | DAKO, M7240 (MIB1), [AB_2142367] | b), c) |
| Ki67 | rat | NA | 1:50 | Thermo Fisher Scientific, 14-5698-82 (SolA15), [AB_10854564] | a), d) |
| NeuN | rabbit | NA | 1:50 | Abcam, ab177487 (EPR12763), [AB_2532109] | a), d) |
| SOX2 | rat | NA | 1:50 | Thermo Fisher Scientific, 14-9811-82 (Btjce), [AB_11219471] | a), d) |
| Tbr1 | rabbit | NA | 1:50 | Abcam, ab31940, [AB_2200219] | a) |
| βIII Tubulin | mouse | NA | 1:1000 | Promega, G712A (5G8), [AB_430874] | b), c) |
| Mouse IgG | donkey | AF 488 | 1:500 | Molecular Probes, A21202, [AB_141607] | |
| Mouse IgG | goat | AF 568 | 1:500 | Molecular Probes, A11004, [AB_2534072] | |
| Rabbit IgG | donkey | AF 546 | 1:500 | Thermo Fisher Scientific, A10040, [AB_2534016] | |
| Rabbit IgG | goat | AF 488 | 1:500 | Thermo Fisher Scientific, A11034, [AB_2576217] | |
| Rat IgG | donkey | AF 549 | 1:500 | Jackson ImmunoResearch Labs, 712-585-153, [AB_2340689] | |

***Supplementary Table S8.***

Immunohistochemistry antibodies used. NA: not applicable (unconjugated). AF: Alexa Fluor. a) cortex, 15pcw; b) cerebellum 12pcw; c) cerebellum 16pcw; d) cerebellum 18pcw.

### C. Supplementary Materials and Methods

***Immunohistochemistry:***

Fetal hindbrain 18pcw was fixed in 4% Paraformaldehyde (PFA) and transferred to methacarn (60% absolute methanol, 30% chloroform, 10% glacial acetic acid) prior to embedding in paraffin wax and subsequent sectioning. Following heat mediated antigen retrieval with citrate buffer pH 6, slides were blocked with 1% BSA overnight at 4˚C and incubated with primary antibody in 1% BSA for 1 hour at room temperature. Slides were incubated with secondary antibodies diluted in 1% BSA at 1:500 45 minutes at room temperature in the dark. Nuclei were stained with DAPI at 1:10.000 in PBS. Slides were Incubated in 0.1% Sudan Black for 20 min at room temperature and mounted with VectaMount AQ aqueous mounting medium. Slides were imaged on the Zeiss Axio Scan.Z1 microscope with the software module ZEN slidescan. Images were further analyzed on QuPath v.0.2.3[17]. Only minor adjustments limited to contrast and brightness to the entire image were performed.

Fetal hindbrain 12pcw and 16pcw were processed and analyzed as described in ref. [18].

Antibodies used are provided in Supplementary Table S8.

***Genetic analysis***

Individuals had previously been diagnosed through Next Generation Sequencing (NGS) panels, exome sequencing, genome sequencing or genomic microarray as part of local diagnostic or research studies for developmental delay, intellectual disability or autism spectrum disorder. All patients had provided informed consent for diagnostic or research testing according to local jurisdiction guidelines and relevant ethical review board requirements.

Where available, methods according to each contributor are detailed.

Patients recruited to the Deciphering Developmental Disorders Study: per ref. [19].

Patients recruited to the 100,000 Genomes Project and Genomics England: per ref. [20].

Patients tested by GeneDx: Using genomic DNA from the proband and parent(s), the exonic regions and flanking splice junctions of the genome were captured using the Clinical Research Exome kit (Agilent Technologies, Santa Clara, CA) or the IDT xGen Exome Research Panel v1.0. Massively parallel (NextGen) sequencing was done on an Illumina system with 100bp or greater paired-end reads. Reads were aligned to human genome build GRCh37/UCSC hg19, and analyzed for sequence variants using a custom-developed analysis tool. Additional sequencing technology and variant interpretation protocol has been previously described[21]. The general assertion criteria for variant classification are publicly available on the GeneDx ClinVar submission page (http://www.ncbi.nlm.nih.gov/clinvar/submitters/26957/).

Patients tested by ARUP Laboratories: Cytogenomic SNP microarray testing was performed for patients 40 and 42 using the CytoScan HD platform (Thermo Fisher Scientific, Inc., Santa Clara, CA, USA).

C4RCD Research Group: PCR-free whole genome sequencing was performed at the research labs at the Translational Genomics Research Institute using the Illumina NovaSeq6000. Emedgene (https://www.emedgene.com/) software was employed for variant analysis.

O.L.B.: Trio sequencing of the proband and the parents was performed by 150bp paired end whole exome sequencing on the Illumina Nextseq 500 with Nextera Rapid Capture Custom Kit. An inhouse bioinformatics pipeline was applied, using bwa alignment and GATK/Picard for variant calling. Variant annotation was performed with Annovar. For variant analysis, the

DDDG2P-gene list (https://decipher.sanger.ac.uk/info/ddg2p) was applied to filter clinically relevant variants using the Filtus tool[22].

A.B.: DNA-laboratory of the Department of Clinical Genetics, Erasmus MC University Medical Center, Rotterdam. Methods available on request.

A.S.: per ref. [23]

B.B.A.d.V: per ref. [24].

K.A., W.B.: per ref. [25].

For additional genetics analyses methods please contact the authors.

### D. Supplementary Information

### Consortia

**C4RCD Research Group**, Center for Rare Childhood Disorders, Translational Genomics Research Institute, Phoenix, Arizona. The following members (listed in alphabetical order) have contributed significantly to this work: Newell Belnap, Anna Bonfitto, Matthew Huentelman, Wayne Jepsen, Vinodh Narayanan, Marcus Naymik, Keri Ramsey, Sampathkumar Rangasamy, Meredith Sanchez-Castillo, and Szabolics Szelinger. This group includes the clinical team and laboratory research team involved in patient enrollment, sample processing, exome sequencing, data processing, preparation of variant annotation files, data analysis, validation of data, and return of research data to families. Candidate genes are identified and discussed at data analysis meetings of the entire group.

**Telethon Undiagnosed Diseases Program:** Vincenzo Nigro[a, b], Annalaura Torella[a, b], Michele Pinelli[a], Manuela Morleo[a], Raffaele Castello[a], Sandro Banfi[a, b], Alessandra Varavallo[a, b], Angelo Selicorni[c], Milena Mariani[c], Silvia Maitz[d], Valeria Capra[e], Andrea Accogli[e], Marcello Scala[e], Vincenzo Leuzzi[f], Anna Commone[f], Francesca Nardecchia[f], Serena Galosi[f], Mario Mastrangelo[f], Donatella Milani[g], Corrado Romano[h], Pinella Failla[h], Donatella Greco[h], Chiara Pantaleoni[i], Claudia Ciaccio[i], Stefano D'Arrigo[i], Nicola Brunetti Pierri[a,j], Giancarlo Parenti[a, j], Gerarda Cappuccio[a,j], Antonietta Coppola[j], Alice Donati[k], Martino Montomoli[k], Teresa Mattina[l], Marcella Zollino[m], Simona Amenta[m], Albina Tummolo[n], Claudia Santoro[o], Anna Grandone[o], Silverio Perrotta[o], Emanuele Miraglia Del Giudice[o], Daniele De Brasi[p], Antonio Varone[p], Gennaro Oliva[q], Margherita Mutarelli[a, r], Angela Peron[s,t]

[a]Telethon Institute of Genetics and Medicine, Pozzuoli, Naples, Italy; [b]Department of Precision Medicine, University of Campania "Luigi Vanvitelli", Naples, Italy; [c]Department of Pediatrics, ASST Lariana Sant'Anna Hospital, San Fermo Della Battaglia, Como, Italy; [d]MBBM Foundation, Monza, Italy; [e]Neuroscience Department, Giannina Gaslini Institute, Genoa, Italy; [f]Department of Human Neuroscience, Sapienza University of Rome, Italy; [g]Pediatric Highly Intensive Care Unit, Fondazione IRCCS Ca' Granda, Ospedale Maggiore Policlinico, Milan, Italy; [h]Oasi Research Institute - IRCCS, Troina, Italy; [i]Developmental Neurology Unit, Fondazione IRCCS Istituto Neurologico Carlo Besta, Milan, Italy; [j]Department of Translational Medicine, Section of Pediatrics, Federico II University, Naples, Italy; [k]Department, A Meyer Children's Hospital, University of Florence, Florence, Italy; [l]Department of Biomedical and Biotechnological Sciences, University of Catania, Catania, Italy; [m]Institute of Genomic Medicine, Catholic University, Gemelli Hospital Foundation, Rome, Italy; [n]Department of Metabolic Diseases, Clinical Genetics and Diabetology, Giovanni XXIII Children's Hospital, Bari, Italy; [o]Pediatric Surgery, Department of Women, Children, General, and Specialist

Surgery, Campania University “Luigi Vanvitelli”, Naples, Italy; [p]Department of Pediatrics, AORN Santobono Pausilipon, Naples, Italy; [q]Institute for High Performance Computing and Networking, National Research Council, Naples, Italy; [r]Institute of Applied Sciences and Intelligent Systems “Eduardo Caianiello” , ISASI, National Research Council, Naples, Italy; [s]Human Pathology and Medical Genetics, ASST Santi Paolo e Carlo, San Paolo Hospital, Milan, Italy; [t]Child Neuropsychiatry Unit - Epilepsy Center, Department of Health Sciences, ASST Santi Paolo e Carlo, San Paolo Hospital, Università Degli Studi di Milano, Milan, Italy.

**University of Washington Center for Mendelian Genomics (UW-CMG)**: Michael J. Bamshad[a,b], Suzanne M. Leal[c], and Deborah A. Nickerson[a], Peter Anderson[a], Tamara J. Bacus[a], Elizabeth E. Blue[a], Kati J. Buckingham[a], Jessica X. Chong[a], Diana Cornejo Sánchez[c], Colleen P. Davis[a], Christian D. Frazar[a], Danielle Giroux[a], William W. Gordon[a], Martha Horike-Pyne[a], Jameson R. Hurless[a], Gail P. Jarvik[a], Eric Johanson[a], J. Thomas Kolar[a], Melissa P. MacMillan[a], Colby T. Marvin[a], Sean McGee[a], Daniel J. McGoldrick[a], Betselote Mekonnen[a], Patrick M. Nielsen[a], Karynne Patterson[a], Benjamin Pelle[a], Aparna Radhakrishnan[a], Matthew A. Richardson[a], Gwendolin T. Roote[a], Erica L. Ryke[a], Isabelle Schrauwen[c], Kathryn M. Shively[a], Joshua D. Smith[a], Monica Tackett[a], Machiko S. Threlkeld[a], Gao Wang[c], Jeffrey M. Weiss[a], Marsha M. Wheeler[a], Qian Yi[a], Jordan E. Zeiger[a], and Xiaohong Zhang[a].
[a]University of Washington, [b]Seattle Children’s Hospital, [c]Columbia University
The full list of current members of University of Washington Center for Mendelian Genomics is available at http://uwcmg.org/docs/Crediting_UW-CMG/UW_CMG_Banner.pdf.

| Reference | Patient ID | Sex | NM 022893.4 | NP 075044.2 (XL isoform)[1] | Inheritrance | Mutation class | Stop or FS disrupting ZNF (I: 378-427; II: 742-823) | Age at diagnosis | Age at last eval | Prenatal history | Prenatal/birth history detail | GW at birth[2] | Congenital malformations |
|---|---|---|---|---|---|---|---|---|---|---|---|---|---|
| HPO terms | | | | | | | | | | | HP:0001197 | | |
| | **P1 (dizygotic twin of P2)** | M | c.12_19dup | p.Gly7AlafsTer9 | assumed germline mosaicisr | PTVa1 | FS -ZNF I&II | 6-10 yrs | 11-15 yrs | normal | twin pregnancy, | 36 | no |
| | **P2 (dizygotic twin of P1)** | M | c.12_19dup | p.Gly7AlafsTer9 | assumed germline mosaicisr | PTVa1 | FS -ZNF I&II | 6-10 yrs | 11-15 yrs | normal | twin pregnancy, | 36 | no |
| | **P3** | F | c.53C>A | p.Ser18Ter | de novo | PTVa1 | stop | NA | 1-5 yrs | normal | | 39 | no |
| Beleford et al., 2020 | **P4** | F | c.148C>T | p.Gln50Ter | de novo | PTVa1 | stop | 1-5 yrs | 1-5 yrs | abnormal | gestational diab | 39 | yes |
| | **P5** | M | c.156_157insCCTG | p.Met53ProfsTer32 | de novo | PTVa2 | FS -ZNF I&II | 1-5 yrs | 1-5 yrs | normal | | 40 | no |
| | **P6** | M | c.263C>A | p.Ser88Ter | de novo | PTVa2 | stop | 1-5 yrs | 6-10 yrs | normal | | 40 | no |
| | **P7** | F | c.286_291delinsA | p.Ser96ThrfsTer13 | de novo | PTVa2 | FS -ZNF I&II | NA | 6-10 yrs | normal | | 40 | no |
| Aldinger et al., 2019 | **P8** | M | c.295del | p.Val99TrpfsTer29 | de novo | PTVa2 | FS -ZNF I&II | NA | 11-15 yrs | normal | | NA | no |
| | **P9** | F | c.363C>A | p.Cys121Ter | NA | PTVa2 | stop | 6-10 yrs | 6-10 yrs | abnormal | maternal foot s | 39 | yes |
| | **P10** | M | c.502dup | p.Ser168LysfsTer69 | not in one parent, other not te | PTVa1 | FS -ZNF I&II | NA | 11-15 yrs | NA | | 33 | no |
| | **P11** | F | c.633_643del | p.Gly212ArgfsTer21 | de novo | PTVb1 | FS -ZNF I&II | 11-15 yrs | 11-15 yrs | normal | | 40 | no |
| | **P12** | M | c.794del | p.Leu265ArgfsTer15 | de novo | PTVb1 | FS -ZNF I&II | 1-5 yrs | 1-5 yrs | abnormal | maternal gestos | 36 | no |
| | **P13** | M | c.794del | p.Leu265ArgfsTer15 | de novo | PTVb1 | FS -ZNF I&II | 16-20 yrs | 16-20 yrs | normal | | 40 | no |
| | **P14** | M | c.952A>T | p.Arg318Ter | de novo | PTVb1 | stop | NA | 6-10 yrs | normal | | 40 | yes |
| | **P15** | M | c.1078dup | p.Leu360ProfsTer212 | de novo | PTVb1 | FS -ZNF I&II | 11-15 yrs | 11-15 yrs | normal | | 37 | no |
| | **P16** | F | c.1078dup | p.Leu360ProfsTer212 | de novo | PTVb1 | FS -ZNF I&II | NA | 6-10 yrs | NA | | 40 | no |
| Dias et al., 2016 | **P17** | F | c.1078dup | p.Leu360ProfsTer212 | de novo | PTVb1 | FS -ZNF I&II | 16-20 yrs | 11-15 yrs | normal | but emergency | 38 | no |
| | **P18** | M | c.1118dup | p.Val374GlyfsTer198 | de novo | PTVb1 | FS -ZNF I&II | 11-15 yrs | 11-15 yrs | normal | | 40 | no |
| | **P19** | M | c.1133C>G | p.Ser378Ter | de novo | PTVb1 | stop | 11-15 yrs | 11-15 yrs | abnormal | oligohydramnios | 33 | no |
| | **P20** | F | c.1287_1288insCACA | p.Lys430HisfsTer143 | de novo | PTVb2 | FS -ZNF II | 1-5 yrs | 6-10 yrs | abnormal | gestational diab | NA | no |
| | **P21** | M | c.1411A>T | p.Lys471Ter | de novo | PTVb2 | stop | 1-5 yrs | 1-5 yrs | normal | | 38 | no |
| | **P22 (sibling of P23)** | F | c.1459G>T | p.Glu487Ter | assumed germline mosaicisr | PTVb2 | stop | 11-15 yrs | NA | normal | | 42 | no |
| | **P23 (sibling of P22)** | F | c.1459G>T | p.Glu487Ter | assumed germline mosaicisr | PTVb2 | stop | 11-15 yrs | NA | normal | | 40 | no |
| Aldinger et al., 2019 | **P24** | M | c.1601_1631del | p.Val534AlafsTer54 | de novo | PTVb2 | FS -ZNF II | NA | 21-25 yrs | normal | | NA | no |
| | **P25** | F | c.1690del | p.Gln564ArgfsTer34 | de novo | PTVb2 | FS -ZNF II | 1-5 yrs | 1-5 yrs | abnormal | maternal hypert | 40 | no |
| | **P26** | M | c.1735_1741del | p.Glu579ThrfsTer17 | de novo | PTVb2 | FS -ZNF II | 6-10 yrs | 6-10 yrs | abnormal | mild ventriculon | 39 | no |
| | **P27** | M | c.1831G>T | p.Glu611Ter | de novo | PTVb2 | stop | 6-10 yrs | 6-10 yrs | normal | | 39 | no |
| | **P28** | F | c.1847dup | p.Leu617ProfsTer18 | de novo | PTVb2 | FS -ZNF II | 1-5 yrs | 1-5 yrs | abnormal | oligohydramnios | 35 | yes |
| | **P29** | M | c.1967_1968dup | p.Ser657ThrfsTer134 | inherited | PTVb2 | FS -ZNF II | 16-20 yrs | 16-20 yrs | NA | | 38 | yes |
| | **P30** | F | c.2035_2036del | p.Ser679GlnfsTer47 | de novo | PTVb2 | FS -ZNF II | 1-5 yrs | 1-5 yrs | normal | | 40 | no |
| | **P31** | M | c.2192_2201dup | p.Ser734ArgfsTer15 | inherited | PTVb2 | FS -ZNF II | 11-15 yrs | 11-15 yrs | abnormal | maternal hypert | 40 | no |
| | **P32** | M | c.55+1G>T | p.(?) | de novo | SPL | | 6-10 yrs | 6-10 yrs | normal | | 38 | no |
| | **P33** | F | c.56-1G>A | p.(?) | de novo | SPL | | 1-5 yrs | 6-10 yrs | abnormal | several materna | 40 | no |
| | **P34** | M | c.385+2T>C | p.(?) | de novo | SPL | | NA | 1-5 yrs | normal | | 40 | no |
| | **P35** | F | c.139A>C | p.Thr47Pro | de novo | MISS | | 6-10 yrs | 6-10 yrs | abnormal | single umbilical | 39 | no |
| | **P36** | M | c.143G>T | p.Cys48Phe | de novo | MISS | | NA | NA | normal | | 40 | no |
| | **P37** | F | c.563A>G | p.His188Arg | de novo | MISS | | 6-10 yrs | 1-5 yrs | normal | | 39 | no |
| | **P38** | M | c.2268T>G | p.Asn756Lys | de novo | MISS | | 16-20 yrs | 16-20 yrs | normal | | 40 | no |
| | **P39** | M | [GRCh38] 2p16.1(60508596-60555113)x1 | | NA | CNV | | 1-5 yrs | 1-5 yrs | normal | | 27 | no |
| | **P40** | F | [GRCh38] 2p16.1(60533641-60561918)x1 | | NA | CNV | | 6-10 yrs | 6-10 yrs | normal | | 40 | no |
| | **P41** | F | [GRCh38] 2p16.1(59833747-60655840)x1 | | de novo | CNV | | 1-5 yrs | 6-10 yrs | normal | | 39 | no |
| | **P42** | M | [GRCh38] 2p16.1(60466611-60575172)x1 | | NA | CNV | | 16-20 yrs | 16-20 yrs | NA | | NA | NA |
| **Previously reported** | | | | | | | | | | | | | |
| Dias et al., 2016 | **Dias_8** | F | c.154C>T | p.Gln52Ter | de novo | PTVa2 | stop | NA | NA | NA | | 39 | no |
| Dias et al., 2016 | **Dias_9** | M | c.193G>T | p.Glu65Ter | de novo | PTVa2 | stop | NA | NA | NA | | 41 | no |
| Korenke et al., 2020 | **Korenke_1** | M | c.271del | p.Glu91ArgfsTer2 | de novo | PTVa2 | FS -ZNF I&II | 13 | 13 | normal | | 40 | no |
| Dias et al., 2016 | **Dias_4** | F | c.529C>T | p.Gln177Ter | de novo | PTVa1 | stop | 8.8 | 8.8 | abnormal | gestational diab | 37 | no |
| Yoshida et al., 2017 | **Yoshida_1** | M | c.577del | p.His193MetfsTer3 | de novo | PTVa1 | FS -ZNF I&II | NA | 15 | NA | | 29 | no |
| Cai et al., 2017 | **Cai_1** | M | c.644C>G | p.Ser215Ter | de novo | PTVb1 | stop | NA | 3.5 | NA | | NA | no |
| Dias et al., 2016 | **Dias_11** | M | c.793dup | p.Leu265ProfsTer3 | NA | PTVb1 | FS -ZNF I&II | NA | NA | NA | | NA | no |
| Soblet et al., 2018 | **Soblet_1** | M | c.1343del | p.Pro448ArgfsTer31 | de novo | PTVb2 | FS -ZNF II | 7 | 7 | normal | | NA | no |
| Dias et al., 2016 | **Dias_10** | F | c.1325del | p.Leu442ProfsTer37 | NA | PTVb2 | FS -ZNF II | NA | NA | NA | | NA | no |
| Wessels et al., 2021 | **Wessels_1** | F | c.1453G>T | p.Glu485Ter | de novo | PTVb2 | stop | 3.8 | 7 | normal | | NA | no |
| Dias et al., 2016 | **Dias_6** | F | c.1540_1544dup | p.Phe515LeufsTer5 | de novo | PTVb2 | FS -ZNF II | NA | 4.4 | NA | | NA | no |
| Dias et al., 2016 | **Dias_7** | F | c.1775_1776insTGGCTCAGCGG | p.Glu593GlyfsTer9 | de novo | PTVb2 | FS -ZNF II | NA | NA | NA | | 39 | no |
| Cai et al., 2017 | **Cai_2** | M | c.1826del | p.Pro609ArgfsTer21 | NA | PTVb2 | FS -ZNF II | NA | 6 | NA | | NA | no |
| Dias et al., 2016 | **Dias_3** | F | c.198C>A | p.His66Gln | de novo | MISS | | NA | 7.75 | normal | | 39 | no |
| Yoshida et al., 2017 | **Yoshida_2** | M | c.2351A>C | p.Lys784Thr | de novo | MISS | | NA | 9 | NA | | 40 | no |
| Balci et al., 2015; Bas | **Balci_1_Basak_3** | F | [GRCh38] 2p16.1(59731285-60607163)x1 | | de novo | CNV | | 3 | 3 | abnormal | increased risk fo | 34 | yes |
| Peter et al., 2014 | **Peter_1** | M | [GRCh38] 2p16.1(60462164-60603356)x1 | | de novo | CNV | | 11 | 11 | normal | | NA | no |
| **Publicly accessible databases** | | | | | | | | | | | | | |
| Decipher_291669 | **Decipher_291669** | F | c.16C>T | p.Gln6Ter | de novo | PTVa1 | stop | NA | 3.6 | NA | | NA | NA |
| ClinVar_VCV0009870 | **ClinVar_VCV000987092.1** | NA | HGVSc=c.55_55+1insT | p.Pro19LeufsTer5 | NA | PTVa1[4] | FS -ZNF I&II | NA | NA | NA | | NA | NA |
| ClinVar_VCV0009873 | **ClinVar_VCV000987335.1** | NA | c.370C>T | p.Gln124Ter | NA | PTVa2 | stop | NA | NA | NA | | NA | yes |
| Decipher_261208 | **Decipher_261208** | M | c.596T>G | p.Leu199Ter | inherited | PTVa1 | stop | NA | 4.3 | NA | | NA | no |
| Decipher_400685 | **Decipher_400685** | F | c.599_602del | p.Glu200AlafsTer7 | de novo | PTVa1 | FS -ZNF I&II | NA | 17 | abnormal | IUGR | NA | no |
| ClinVar_VCV0009861 | **ClinVar_VCV000986112.1** | NA | c.794del | p.Leu265ArgfsTer15 | NA | PTVb1 | FS -ZNF I&II | NA | NA | NA | | NA | NA |
| ClinVar_VCV0009870 | **ClinVar_VCV000987079.1** | NA | c.1078del | p.Leu360SerfsTer61 | NA | PTVb1 | FS -ZNF I&II | NA | NA | NA | | NA | NA |
| ClinVar_VCV0009868 | **ClinVar_VCV000986806.1** | NA | c.1555_1556del | p.Leu519GlyfsTer52 | NA | PTVb2 | FS -ZNF II | NA | NA | NA | | NA | NA |
| ClinVar_VCV0009853 | **ClinVar_VCV000985399.1** | NA | c.2194dup | p.Arg732LysfsTer14 | NA | PTVb2 | FS -ZNF II | NA | NA | NA | | NA | yes |
| ClinVar_VCV0009859 | **ClinVar_VCV000985925.1** | NA | c.55+1G>A | p.(?) | NA | SPL | | NA | NA | NA | | NA | NA |
| ClinVar_VCV0009870 | **ClinVar_VCV000987091.1** | NA | c.55+1_55+2insCCCAA | p.(?) | NA | SPL | | NA | NA | NA | | NA | NA |
| ClinVar_VCV0009870 | **ClinVar_VCV000987090.1** | NA | c.55+5del | p.(?) | NA | SPL | | NA | NA | NA | | NA | NA |
| ClinVar_VCV0009870 | **ClinVar_VCV000987093.1** | NA | c.55C>T | p.Pro19Ser | NA | MISS[5] | | NA | NA | NA | | NA | NA |
| Decipher_300584 | **Decipher_300584** | M | [GRCh38] 2p16.1(60452463-60629834)x1 | | de novo | CNV | | NA | 7.4 | NA | | NA | yes |
| Decipher_300585 | **Decipher_300585** | M | [GRCh38] 2p16.1(60452463-60629834)x1 | | de novo | CNV | | NA | 7.4 | NA | | NA | yes |
| Decipher_305786 | **Decipher_305786** | F | [GRCh38] 2p16.1(60214168-60469017)x1 | | de novo | CNV | | NA | 3.7 | NA | | NA | no |

***Supplementary Table S1.*** Detailed clinical data of individuals reported.

[1] see Supplementary Table S2 for other isoforms

[2] when gestational age at birth was recorded as "at term", that was converted to 40 gestational weeks

[3] Thick eyebrows or long eyelashes or increased body hair

[4] frameshift + splice site disruption (SpliceAI, donor_gain and donor_loss)

[5] missense and potential splice site disruption (SpliceAI)

[6] raw number was not available; reported as is in the publication or in the publicly available database

Abbreviations: HPO: human phenotype ontology; db: database; NA: not available; eval: evaluation; IVF: in vitro fertilization; IUGR: intrauterine growth restriction; GW: gestational week; HC: head circumference; ACTH: Adrenocorticotropic hormone; abn: abnormalities; IV Ig: intravenous immunoglobulins; HbF: fetal hemoglobin; CMA: chromosomal microarray; WES: whole exome sequencing; seq: sequencing; mDNA: mitochondrial DNA; NGS: next generation sequencing; VUS: variant of unknown clinical significance; SMA: spinal muscolar atrophy; CK: creatine kinase.

| Congenital malformation detail | HC at birth (cm) | HC at birth (SD, UKWHO) | Age at last observation HC | HC at last observation (cm) | HC at last observation (SD, UKWHO) | Age at last observation height | Height at last observation (cm) | Height at last observation (SD, | Age at last observation weight | Weight at last observation (kg) | Weight at last observation (SD, | Hirsutism[3] | Epicanthus | Wide nose | Malar flattening | Full cheeks |
|---|---|---|---|---|---|---|---|---|---|---|---|---|---|---|---|---|
| | | HP:0011451 | | | HP:0005484<br>HP:0000253 | | | HP:0004322 | | | | HP:0000998<br>HP:0011229<br>HP:0000527 | HP:0000286 | HP:0000445<br>HP:0012810<br>HP:0000431<br>HP:0000414<br>HP:0000455 | HP:0000272 | HP:0000293 |
| | NA | NA | 11-15 yrs | 51.5 | -2.11240385 | 11-15 yrs | 139 | -0.91365456 | 11-15 yrs | 29.7 | -1.20128856 | no | no | yes | yes | no |
| | NA | NA | 11-15 yrs | 51.5 | -2.11240385 | 11-15 yrs | 139 | -0.91365456 | 11-15 yrs | 29.8 | -1.17804533 | no | no | yes | yes | no |
| | 36 | 2.03263425 | 1-5 yrs | 47.3 | -1.02382695 | 1-5 yrs | 101 | 0.993048673 | 1-5 yrs | 14.9 | 0.251112163 | no | no | no | yes | yes |
| bilateral post-a | NA | NA | 1-5 yrs | 46 | -1.77678414 | 1-5 yrs | 95.5 | -1.65554971 | 1-5 yrs | 14.5 | -1.01571998 | no | no | no | yes | yes |
| | 35 | 0.07685577 | 1-5 yrs | 46.5 | -1.29 | 1-5 yrs | 85 | -0.81 | 1-5 yrs | 10.9 | -0.95 | no | no | no | no | no |
| | NA | NA | 1-5 yrs | 48.3 | -0.8177286 | 1-5 yrs | 94.3 | -0.4811296 | 1-5 yrs | 16.1 | 0.950027745 | no | no | yes | yes | no |
| | 32.5 | -1.56328614 | 6-10 yrs | 47.5 | -3.90137726 | 6-10 yrs | 116 | 0.138122122 | 6-10 yrs | 22.5 | 0.637892353 | no | yes | yes | yes | yes |
| | NA (rep | normal[6] | 6-10 yrs | 51 | -1.8726551 | 6-10 yrs | 126.8 | -0.45459731 | 6-10 yrs | 29.8 | 0.784273177 | no | no | no | yes | yes |
| small umbilica | NA | NA | 1-5 yrs | 48 | -0.49683285 | 1-5 yrs | 94 | -0.68653667 | 1-5 yrs | 15.1 | 0.411444541 | no | yes | no | no | yes |
| | NA | NA | 11-15 yrs | 54.4 | -0.59391552 | NA | NA | NA | 11-15 yrs | 41.6 | 0.033459213 | yes | yes | no | yes | no |
| | NA | NA | 11-15 yrs | 52 | -2.05328069 | 11-15 yrs | 142 | -2.24241563 | 11-15 yrs | 45.2 | -0.27834902 | no | no | no | no | no |
| | 30.5 | -1.64827001 | 1-5 yrs | 46 | -2.31716706 | 1-5 yrs | 95 | 0.144660154 | 1-5 yrs | 12.5 | -0.90912194 | yes | no | yes | no | no |
| | NA | NA | 16-20 yrs | 55 | -1.1170359 | 16-20 yrs | 176 | 0.036255692 | 16-20 yrs | 58.2 | -0.65723879 | no | yes | yes | yes | yes |
| cleft palate, m | NA | NA | 6-10 yrs | 49 | -3.14094816 | 6-10 yrs | 130 | -1.53945597 | 6-10 yrs | 29 | -0.66055116 | no | no | yes | no | yes |
| | 34 | 0.50636953 | 6-10 yrs | 48 | -4.12647975 | 6-10 yrs | 123 | -2.80149586 | 6-10 yrs | 30.8 | -0.42152411 | no | no | yes | yes | yes |
| | NA | NA | 6-10 yrs | 50 | -2.99190095 | 6-10 yrs | 138.3 | -0.10866351 | 6-10 yrs | 26.6 | -1.21515479 | no | no | no | no | no |
| | NA | NA | 11-15 yrs | 53.5 | -0.57274224 | 11-15 yrs | 151 | 0.494665926 | 11-15 yrs | 53.6 | 1.600995972 | no | no | no | yes | no |
| | NA | NA | 11-15 yrs | 54.2 | -1.15018488 | 11-15 yrs | 159.3 | -0.95453532 | 11-15 yrs | 47.6 | -0.66091324 | no | yes | no | yes | no |
| | 34 | 2.18451193 | NA | NA | NA | NA | NA | NA | NA | NA | NA | no | yes | no | yes | no |
| | NA | NA | NA | NA (1.5th c) | -2.17 | NA | NA (50th c) | 0 | NA | NA (5th c) | -1.64 | no | yes | yes | no | yes |
| | NA | NA | 1-5 yrs | 45 | -2.96344149 | 1-5 yrs | 88.4 | -1.48741272 | 1-5 yrs | 12.9 | -0.52183078 | no | no | yes | no | no |
| | 35 | -0.01401506 | 1-5 yrs | 49.5 | -1.39286214 | 6-10 yrs | 118 | -0.40448823 | 6-10 yrs | 23 | 0.164079827 | yes | yes | yes | no | yes |
| | 34 | -0.12504985 | 6-10 yrs | 52 | -0.43103916 | 6-10 yrs | 121 | 0.183094579 | NA | NA | NA | no | no | yes | yes | yes |
| | NA | NA | NA | NA | NA | NA | NA | NA | NA | NA | NA | no | no | no | no | no |
| | NA | NA | 1-5 yrs | 49 | -1.88900773 | 1-5 yrs | 104 | 0.166806381 | 1-5 yrs | 16 | -0.34514974 | no | no | yes | no | yes |
| | 35 | 0.4819678 | 1-5 yrs | 50.5 | -1.46385717 | 1-5 yrs | 105 | -0.86649293 | 1-5 yrs | 16.8 | -0.78014044 | yes | yes | yes | no | no |
| | NA | NA | 6-10 yrs | 50.3 | -2.48735219 | 6-10 yrs | 135 | 0.073020178 | 6-10 yrs | 29.1 | -0.00065451 | no | no | no | no | no |
| congenital ste | 28.5 | -2.54817256 | 1-5 yrs | 44.5 | -2.69213793 | 1-5 yrs | 87.6 | -1.55569625 | 1-5 yrs | 9.6 | -2.75232209 | yes | yes | yes | yes | yes |
| craniosynostos | NA | NA | 16-20 yrs | 53 | -2.48737831 | 16-20 yrs | 177.5 | 0.058494365 | 16-20 yrs | 62 | -0.55588861 | yes | no | yes | yes | no |
| | NA | NA | 1-5 yrs | 47 | -3.58851074 | NA | NA | NA | NA | NA (9th c) | -1.34 | no | yes | yes | yes | yes |
| | 34.5 | -0.35351589 | 11-15 yrs | 54 | -1.33331212 | 11-15 yrs | 165 | -0.48476178 | 11-15 yrs | 72.2 | 1.431584363 | yes | no | no | no | no |
| | 34 | 0.08307767 | 6-10 yrs | 49.5 | -2.54679306 | 6-10 yrs | 127 | 1.297219095 | 6-10 yrs | 20.5 | -0.710678 | no | no | no | yes | yes |
| | NA | NA | NA | NA | NA | NA | NA | NA | NA | NA | NA | no | no | no | no | no |
| | 37 | 1.79834241 | NA | NA | NA | NA | NA | NA | NA | NA | NA | yes | no | no | no | no |
| | NA | NA | 1-5 yrs | 47.9 | 0.174441091 | 6-10 yrs | 132.7 | -0.02053827 | 6-10 yrs | 28.7 | -0.0364012 | yes | no | no | yes | yes |
| | NA | NA | <1 yrs | 39 | -0.61282771 | <1 yrs | 58 | -0.83869146 | <1 yrs | 5.8 | -0.165191 | no | no | yes | yes | yes |
| | NA | NA | 1-5 yrs | 51 | -0.48006597 | 1-5 yrs | 105 | -0.54576836 | NA | NA | NA | no | no | no | no | no |
| | NA | NA | 16-20 yrs | 56 | 0 | 16-20 yrs | 166 | -1.61697989 | 16-20 yrs | 52 | -2.26403622 | yes | no | yes | yes | no |
| | NA | NA | NA | NA | NA | NA | NA | NA | NA | NA | NA | no | yes | no | no | no |
| | NA | NA | 1-5 yrs | 48.9 | 0.206177955 | 1-5 yrs | 114.3 | 0.302376349 | 1-5 yrs | 20.75 | 0.404902027 | no | no | yes | yes | yes |
| | NA | -0.67 | 6-10 yrs | 50 | -2.23788244 | 6-10 yrs | 119.5 | -0.6273405 | 6-10 yrs | 20.9 | -0.84856406 | no | no | no | no | yes |
| | NA | NA | NA | NA | NA | NA | NA | NA | NA | NA | NA | NA | NA | NA | NA | NA |
| | 35 | 1.11979272 | 12.8 | 53 | -1.19436308 | 12.8 | 161.5 | 1.039587892 | 12.8 | 55.7 | 1.239807562 | no | no | yes | no | no |
| | 35 | -0.34880959 | 12.1 | 54 | -0.7117063 | 12.1 | 163.5 | 2.008525575 | 12.1 | 45.5 | 0.874335521 | no | yes | yes | yes | no |
| | 35 | 0.07685577 | NA | NA (<3rd c) | <3rd centile[6] | NA | NA | NA | NA | NA | NA | NA | NA | NA | NA | NA |
| | NA | NA | 8.8 | 49.2 | -3.33217142 | 8.8 | 131.5 | -0.04255933 | 8.8 | 26.9 | -0.29419678 | no | no | no | yes | no |
| | NA | NA | 15 | 51.3 | -2.94618967 | 15 | 151 | -2.20726559 | NA | NA | NA | no | yes | no | yes | no |
| | NA | NA | 3.5 | 43.5 | -4.41793312 | 3.5 | 87 | -3.24115156 | 3.5 | 12 | -2.00342184 | no | no | no | yes | yes |
| | NA | NA | NA | NA | NA | NA | NA | NA | NA | NA | NA | NA | NA | NA | NA | NA |
| | 33.5 | -0.32 | 7 | 50 | -2.27399662 | NA | NA (normal) | normal[6] | NA | NA (normal) | normal[6] | no | yes | yes | no | no |
| | NA | NA | NA | NA | NA | NA | NA | NA | NA | NA | NA | NA | NA | NA | NA | NA |
| | NA | NA | 0.8 | 41.5 | -1.91431532 | NA | NA (normal) | normal[6] | NA | NA (normal) | normal[6] | no | yes | no | yes | no |
| | NA | NA | NA | NA (<3rd c) | <3rd centile[6] | NA | NA (<3rd c) | <3rd centile[6] | NA | NA | NA | NA | NA | NA | NA | NA |
| | NA | NA | 2.75 | 46 | -1.58967514 | 2.75 | 92 | -0.2515826 | 2.75 | 13.6 | 0.18406951 | yes | yes | yes | no | yes |
| | NA | NA | NA | NA | NA | NA | NA | NA | NA | NA | NA | NA | NA | NA | NA | NA |
| | NA | NA | NA | NA (<3rd c) | <3rd centile[6] | NA | NA (9th c) | -1.34 | NA | NA | -0.65 | no | no | yes | yes | no |
| | NA | NA | 9 | 50 | -2.63464888 | 9 | 127 | -1.08294295 | NA | NA | NA | no | yes | no | yes | no |
| atrial septal de | 31.75 | 0.36570209 | 3 | 47.5 | -0.71491864 | 3 | 94 | -0.27614638 | 3 | 14.1 | 0.137912406 | no | no | no | yes | yes |
| | NA | NA | NA | NA (normal) | normal[6] | NA | NA (normal) | normal[6] | NA | NA (normal) | normal[6] | NA | NA | NA | NA | NA |
| | NA | NA | NA | NA | NA | NA | NA | NA | NA | NA | NA | NA | NA | NA | NA | NA |
| | NA | NA | NA | NA | NA | NA | NA | NA | NA | NA | NA | NA | NA | NA | NA | NA |
| rudimentary p | NA (co | <3rd centile[6] | NA | NA (microce | <3rd centile[6] | NA | NA | NA | NA | NA | NA | NA | yes | NA | NA | NA |
| | NA | NA | NA | NA | NA | NA | NA | NA | NA | NA | NA | NA | yes | NA | yes | NA |
| | NA | NA | NA | NA | NA | NA | NA | NA | NA | NA | NA | NA | NA | NA | NA | NA |
| | NA | NA | NA | NA (microce | <3rd centile[6] | NA | NA | NA | NA | NA | NA | NA | NA | NA | NA | NA |
| | NA | NA | NA | NA | NA | NA | NA (short sta | <3rd centile[6] | NA | NA (obesity) | obesity[6] | NA | NA | NA | NA | NA |
| | NA | NA | NA | NA | NA | NA | NA | NA | NA | NA | NA | NA | NA | NA | NA | NA |
| trigonocephaly | NA | NA | NA | NA | NA | NA | NA | NA | NA | NA | NA | NA | NA | NA | NA | NA |
| | NA | NA | NA | NA | NA | NA | NA (short sta | <3rd centile[6] | NA | NA | NA | NA | NA | NA | NA | NA |
| | NA | NA | NA | NA | NA | NA | NA | NA | NA | NA | NA | NA | NA | NA | NA | NA |
| | NA | NA | NA | NA | NA | NA | NA | NA | NA | NA | NA | NA | NA | NA | NA | NA |
| | NA | NA | NA | NA | NA | NA | NA | NA | NA | NA | NA | NA | NA | NA | NA | NA |
| polydactyly | NA | NA | NA | NA | NA | NA | NA | NA | NA | NA | NA | NA | yes | NA | NA | NA |
| polydactyly | NA | NA | NA | NA | NA | NA | NA | NA | NA | NA | NA | NA | NA | NA | NA | NA |
| | NA | NA | NA | NA | NA | NA | NA | NA | NA | NA | NA | yes | NA | NA | NA | NA |

| Thin vermillion of the upper lip | Thick or everted vermillion of the lower lip | Cleft or high palate | Ear abnormalities | DD/ID | DD/ID severity | Language development | Absent speech | Dysarthria or speech apraxia | Gross motor development | Fine motor development | Autism spectrum disorder | Behavior | Repetitive behavior or stereotypies | Anxiety | ADHD |
|---|---|---|---|---|---|---|---|---|---|---|---|---|---|---|---|
| HP:0000219 | HP:0000179 HP:0000232 | HP:0000174 HP:0000175 | HP:0000356 HP:0000385 HP:0009907 HP:0000396 HP:0030026 | HP:0001263 HP:0012758 HP:0001249 | HP:0001256 HP:0002342 HP:0010864 | HP:0000750 | HP:0001344 | HP:0001260 HP:0011098 | HP:0002194 | HP:0010862 | HP:0000717 HP:0000729 | HP:0000708 | HP:0008762 HP:0000733 | HP:0000739 | HP:0007018 |
| no | yes | no | yes | yes | mild | delayed | | yes | delayed | delayed | no | abnormal | no | no | yes |
| no | no | yes | no | yes | mild | delayed | | yes | delayed | delayed | no | abnormal | no | no | yes |
| yes | yes | no | no | yes | moderate | delayed | | | delayed | delayed | no | normal | no | no | no |
| no | no | no | yes | yes | moderate | delayed | yes | | delayed | NA | yes | normal | no | no | no |
| no | no | no | no | yes | NA | delayed | | | delayed | delayed | no | normal | no | no | no |
| no | no | no | yes | yes | NA | delayed | | yes | delayed | delayed | no | abnormal | yes | no | yes |
| no | yes | no | no | yes | NA | delayed | yes | | delayed | NA | no | abnormal | yes | no | yes |
| yes | yes | no | yes | yes | NA | delayed | | | delayed | delayed | NA | abnormal | yes | no | no |
| no | no | no | no | yes | moderate | delayed | | | delayed | delayed | no | abnormal | no | yes | no |
| no | no | no | yes | yes | severe | delayed | yes | | delayed | NA | yes | normal | no | no | no |
| no | yes | no | yes | yes | severe | delayed | | | delayed | delayed | yes | abnormal | no | no | no |
| yes | no | yes | no | yes | severe | delayed | yes | | delayed | NA | no | abnormal | no | no | yes |
| no | yes | no | yes | yes | mild | delayed | | | delayed | delayed | no | abnormal | yes | no | yes |
| yes | no | yes | yes | yes | NA | delayed | | yes | normal | normal | no | abnormal | no | no | yes |
| yes | yes | no | yes | yes | severe | delayed | yes | | delayed | delayed | no | abnormal | yes | no | no |
| no | yes | no | no | yes | moderate | delayed | | | delayed | delayed | yes | abnormal | yes | no | no |
| no | yes | no | yes | yes | moderate | delayed | | | delayed | NA | no | abnormal | yes | no | no |
| no | no | no | yes | yes | moderate | delayed | | | delayed | delayed | yes | abnormal | yes | no | no |
| no | yes | no | yes | no | IQ 93 | normal but dyslexia and dysgraphia | | | NA | delayed | yes (Asperger) | abnormal | no | no | yes |
| no | yes | no | yes | yes | NA | delayed | | | delayed | delayed | no | normal | no | no | no |
| no | yes | no | yes | yes | NA | delayed | | | delayed | NA | no | abnormal | no | no | no |
| yes | yes | no | no | yes | moderate | delayed | | | delayed | NA | no | normal | no | no | no |
| no | yes | no | no | yes | severe | delayed | yes | | delayed | NA | yes | abnormal | yes | no | no |
| no | yes | no | no | yes | NA | delayed | | | delayed | delayed | NA | abnormal | no | yes | no |
| yes | yes | no | yes | yes | severe | delayed | yes | | delayed | delayed | yes | abnormal | no | no | no |
| yes | no | no | yes | yes | moderate | delayed | | | delayed | normal | yes | abnormal | no | no | yes |
| no | no | no | no | yes | mild | normal | | | delayed | NA | no | abnormal | yes | no | no |
| yes | no | no | yes | NA | NA | NA | | | NA | NA | NA | NA | NA | NA | NA |
| no | yes | yes | yes | no | IQ 80 | normal but speech therapy in early childhoo | | | normal | NA | yes | abnormal | no | no | yes |
| no | yes | no | yes | yes | moderate | delayed | | | delayed | delayed | no | normal | no | no | no |
| yes | no | yes | no | yes | mild | delayed | | | delayed | delayed | no | normal | no | no | no |
| yes | no | no | yes | yes | moderate | delayed | | | delayed | NA | no | normal | no | no | no |
| no | no | no | no | yes | moderate | delayed | | yes | delayed | NA | NA | abnormal | no | yes | no |
| yes | no | no | no | yes | severe | delayed | yes | | delayed | delayed | NA | NA | NA | NA | NA |
| no | yes | no | yes | yes | moderate | delayed | | | delayed | delayed | no | abnormal | yes | no | no |
| no | yes | no | no | yes | moderate | delayed | | | delayed | NA | yes | abnormal | yes | no | no |
| no | no | no | no | yes | NA | delayed | | | delayed | NA | yes | normal | no | no | no |
| yes | no | yes | yes | yes | mild | NA | | | delayed | delayed | no | abnormal | yes | yes | no |
| yes | no | no | no | yes | NA | delayed | | | delayed | NA | no | normal | no | no | no |
| no | no | no | no | yes | mild | delayed | | yes | delayed | delayed | no | normal | no | no | no |
| no | yes | no | yes | yes | NA | delayed | | yes | delayed | delayed | no | normal | no | no | no |
| NA | NA | NA | NA | NA | NA | NA | | | NA | NA | yes | NA | NA | NA | NA |
| yes | yes | yes | no | yes | moderate | delayed | | | delayed | NA | no | abnormal | no | yes | no |
| yes | yes | no | yes | yes | moderate | delayed | | | delayed | NA | no | normal | no | no | no |
| yes | NA | NA | NA | yes | NA | delayed | | | delayed | NA | yes | NA | NA | NA | NA |
| yes | no | yes | yes | yes | moderate | delayed | | | delayed | delayed | no | abnormal | yes | no | no |
| no | yes | no | yes | yes | severe | delayed | | | delayed | NA | NA | NA | NA | NA | NA |
| no | yes | yes | no | yes | severe | delayed | | yes | NA | NA | NA | NA | NA | NA | NA |
| NA | NA | NA | NA | NA | NA | NA | | | NA | NA | yes | NA | NA | NA | NA |
| no | yes | no | yes | yes | moderate | delayed | | yes | delayed | delayed | no | normal | no | no | no |
| NA | NA | NA | NA | yes | severe | delayed | | | delayed | NA | yes | abnormal | no | no | yes |
| yes | no | no | yes | yes | moderate | delayed | yes | | delayed | NA | no | abnormal | yes | no | no |
| NA | NA | NA | NA | yes | NA | delayed | | | NA | NA | NA | NA | NA | NA | NA |
| no | yes | no | yes | yes | moderate | delayed | | | NA | NA | no | normal | no | no | no |
| NA | NA | NA | NA | yes | NA | delayed | | | delayed | NA | yes | NA | NA | NA | NA |
| yes | no | no | yes | yes | moderate | delayed | | | delayed | delayed | no | abnormal | yes | no | no |
| yes | no | no | yes | yes | profound | delayed | yes | | delayed | NA | NA | NA | NA | NA | NA |
| yes | yes | no | yes | yes | NA | delayed | | | delayed | delayed | no | normal | no | no | no |
| NA | NA | NA | NA | yes | mild | delayed | | yes | delayed | NA | no | abnormal | no | no | yes |
| NA | NA | NA | NA | yes | moderate | NA | | | NA | NA | NA | NA | NA | NA | NA |
| NA | NA | NA | NA | yes | NA | NA | | | NA | NA | NA | NA | NA | NA | NA |
| NA | NA | NA | NA | yes | NA | NA | | | NA | NA | NA | NA | NA | NA | NA |
| NA | yes | NA | NA | NA | specific learnir | NA | | | NA | NA | NA | abnormal | yes | no | no |
| NA | NA | NA | NA | NA | specific learnir | delayed | | yes | NA | NA | NA | NA | NA | NA | NA |
| NA | NA | NA | NA | yes | NA | NA | | | NA | NA | NA | NA | NA | NA | NA |
| NA | NA | NA | NA | yes | mild | NA | | | NA | NA | NA | NA | NA | NA | NA |
| NA | NA | NA | NA | yes | NA | NA | | | NA | NA | NA | NA | NA | NA | NA |
| NA | NA | NA | NA | yes | NA | NA | | | NA | NA | NA | NA | NA | NA | NA |
| NA | NA | NA | NA | yes | NA | delayed | | yes | NA | NA | NA | NA | NA | NA | NA |
| NA | NA | NA | NA | yes | NA | NA | | | NA | NA | NA | NA | NA | NA | NA |
| NA | NA | NA | NA | yes | NA | NA | | | NA | NA | NA | NA | NA | NA | NA |
| NA | NA | NA | NA | yes | NA | NA | | | NA | NA | NA | NA | NA | NA | NA |
| NA | NA | NA | NA | yes | severe | NA | | | NA | NA | NA | NA | NA | NA | NA |
| NA | NA | NA | NA | yes | moderate | NA | | | NA | NA | NA | NA | NA | NA | NA |
| yes | NA | NA | NA | yes | moderate | NA | | | NA | NA | NA | NA | NA | NA | NA |

| Aggressiveness | Sleep disturbance | Seizures | Seizure type | Seizure frequency | EEG | Anti-seizure medications | Drug-resistance | Tone | Ataxia, broad based gait or spastic paraplegia | Brain MRI | Posterior fossa abn | Brain stem abn | Corpus callosum abn | Cortical malformations | Strabismus |
|---|---|---|---|---|---|---|---|---|---|---|---|---|---|---|---|
| HP:0000718 HP:0025160 | HP:0002360 | HP:0001250 | HP:0007359 HP:0002197 HP:0002069 HP:0032794 | | HP:0002353 | | | HP:0001252 | HP:0002066 HP:0002136 HP:0001258 | HP:0410263 HP:0011282 | HP:0000932 HP:0006817 | HP:0002363 | HP:0001273 HP:0002079 | | HP:0000486 |
| yes | no | no | | | NA | | | hypotonia | no | normal | no | no | no | no | yes |
| no | no | no | | | NA | | | hypotonia | no | abnormal | yes | yes | no | no | yes |
| no | no | no | | | abnormal | no | | hypotonia | no | abnormal | yes | yes | yes | no | yes |
| no | no | no | | | NA | | | hypotonia | no | abnormal | no | yes | no | no | yes |
| no | no | no | | | NA | | | hypotonia | no | abnormal | yes | yes | yes | no | no |
| yes | no | no | | | NA | | | hypotonia | NA | abnormal | yes | no | no | no | yes |
| yes | yes | no | | | NA | | | hypotonia | no | normal | no | no | no | no | yes |
| yes | no | no | | | NA | | | hypotonia | yes | abnormal | yes | yes | no | no | NA |
| no | yes | no | | | NA | | | normal | no | normal | no | no | no | no | yes |
| no | no | yes | generalized to | every 2 month | abnormal | valproic acid, c | yes | hypotonia | yes | normal | no | no | no | no | no |
| yes | no | no | | | abnormal | levetiracetam | | hypotonia | no | abnormal | yes | yes | yes | no | yes |
| no | no | no | | | normal | | | hypotonia | yes | normal | no | no | no | no | NA |
| yes | no | single seizure | tonic-clonic | single seizure | NA | no | | hypotonia | no | abnormal | yes | no | no | no | yes |
| no | no | yes | myoclonic | NA | abnormal | valproic acid | no | normal | no | normal | no | no | no | no | no |
| yes | yes | yes | focal, complex | one every few | abnormal | zonisamide, ph | yes | hypotonia | yes | abnormal | yes | yes | yes | no | yes |
| yes | yes | no | | | normal | | | hypotonia | NA | normal | no | no | no | no | yes |
| yes | yes | no | | | normal | | | normal | yes | normal | no | no | no | no | no |
| yes | yes | no | | | NA | | | hypotonia | NA | normal | no | no | no | no | yes |
| no | yes | NA | | | NA | | | NA | NA | NA | NA | NA | NA | NA | NA |
| no | no | no | | | NA | | | hypotonia | no | normal | no | no | no | no | NA |
| yes | no | no | | | NA | | | normal | NA | normal | no | no | no | no | yes |
| no | no | no | | | NA | | | normal | no | normal | no | no | no | no | yes |
| yes | yes | no | | | abnormal | no | | normal | no | normal | no | no | no | no | no |
| no | no | NA | | | NA | | | NA | NA | abnormal | yes | yes | no | no | NA |
| yes | no | yes | myoclonic | NA | NA | NA | NA | hypotonia | no | normal | no | no | no | no | yes |
| yes | yes | yes | generalized to | daily | abnormal | valproic acid | NA | normal | no | abnormal | no | no | no | no | yes |
| no | yes | yes | generalized | NA | abnormal | levetiracetam, | no | hypotonia | NA | abnormal | no | no | no | no | no |
| NA | NA | no | | | NA | | | normal | NA | abnormal (on | yes | NA | no | no | NA |
| no | no | no | | | NA | | | NA | NA | abnormal | no | no | no | no | no |
| no | no | no | | | NA | | | NA | yes | abnormal | no | no | no | no | yes |
| no | no | no | | | normal | | | normal | no | abnormal | no | no | no | yes | no |
| no | no | no | | | NA | | | normal | yes | normal | no | no | no | no | no |
| no | no | no | | | NA | | | hypotonia | yes | normal | no | no | no | no | no |
| NA | NA | no | | | NA | | | hypotonia | NA | abnormal | yes | yes | yes | no | no |
| no | no | no | | | NA | | | normal | yes | abnormal | no | no | no | no | yes |
| no | yes | yes | absence | NA | normal | NA | NA | hypotonia | yes | abnormal | yes | yes | no | no | yes |
| no | no | no | | | NA | | | normal | yes | NA | NA | NA | NA | NA | no |
| no | no | single seizure | NA | single seizure | NA | no | | normal | yes | abnormal | no | no | yes | no | yes |
| no | no | no | | | NA | | | hypotonia | NA | NA | NA | NA | NA | NA | no |
| no | no | yes | focal, staring s | one every few | abnormal | oxcarbazepine | no | hypotonia | yes | abnormal | no | yes | no | yes | yes |
| no | no | no | | | NA | | | hypotonia | NA | abnormal | yes | yes | yes | no | no |
| NA | NA | NA | | | NA | | | NA | NA | NA | NA | NA | NA | NA | NA |
| no | no | no | | | NA | | | NA | NA | abnormal | yes | no | no | no | yes |
| no | no | no | | | NA | | | NA | yes | abnormal | no | no | yes | no | yes |
| NA | yes | yes | myoclonic asta | NA | abnormal | valproic acid, e | no | hypotonia | NA | abnormal | yes | no | no | no | no |
| yes | yes | no | | | NA | | | hypotonia | NA | abnormal | yes | yes | no | no | yes |
| NA | yes | yes | Lennox-Gastau | daily | NA | valproic acid, c | yes | NA | NA | normal | no | no | no | no | yes |
| NA | NA | no | | | NA | | | NA | NA | abnormal | no | yes | no | no | yes |
| NA | NA | NA | | | NA | | | NA | NA | NA | NA | NA | NA | NA | no |
| no | no | no | | | normal | | | hypotonia | NA | normal | no | no | no | no | no |
| no | no | NA | | | NA | | | NA | NA | NA | NA | NA | NA | NA | no |
| no | no | no | | | NA | | | normal | yes | normal | no | no | no | no | yes |
| NA | NA | no | | | NA | | | NA | NA | NA | NA | NA | NA | NA | no |
| no | no | no | | | NA | | | NA | NA | NA | NA | NA | NA | NA | yes |
| NA | NA | yes | NA | NA | NA | NA | NA | NA | NA | NA | NA | NA | NA | NA | no |
| yes | yes | no | | | NA | | | hypotonia | NA | NA | NA | NA | NA | NA | yes |
| NA | NA | yes | West syndrom | NA | NA | valproic acid, A | yes | NA | NA | abnormal | no | no | yes | no | yes |
| no | no | no | | | NA | | | hypotonia | NA | abnormal | yes | yes | no | no | NA |
| no | no | no | | | NA | | | hypotonia | NA | NA | NA | NA | NA | NA | NA |
| NA | NA | NA | | | NA | | | NA | NA | NA | NA | NA | NA | NA | NA |
| NA | NA | NA | | | NA | | | NA | yes | NA | NA | NA | NA | NA | NA |
| NA | NA | NA | | | NA | | | hypotonia | NA | abnormal | yes | no | NA | NA | NA |
| no | no | NA | | | NA | | | NA | NA | NA | NA | NA | NA | NA | NA |
| NA | yes | NA | | | NA | | | NA | NA | NA | NA | NA | NA | NA | NA |
| NA | NA | NA | | | NA | | | NA | NA | NA | NA | NA | NA | NA | NA |
| NA | NA | yes | spasms | NA | NA | NA | NA | NA | NA | NA | NA | NA | NA | NA | NA |
| NA | NA | NA | | | NA | | | NA | NA | NA | NA | NA | NA | NA | NA |
| NA | NA | NA | | | NA | | | hypotonia | NA | NA | NA | NA | NA | NA | NA |
| NA | NA | NA | | | NA | | | hypotonia | NA | NA | NA | NA | NA | NA | NA |
| NA | NA | NA | | | NA | | | NA | yes | NA | NA | NA | NA | NA | NA |
| NA | NA | NA | | | NA | | | NA | yes | NA | NA | NA | NA | NA | NA |
| NA | NA | NA | | | NA | | | NA | yes | NA | NA | NA | NA | NA | NA |
| NA | NA | NA | | | NA | | | NA | NA | NA | NA | NA | NA | NA | NA |
| NA | NA | NA | | | NA | | | NA | NA | NA | NA | NA | NA | NA | NA |
| NA | NA | NA | | | NA | | | NA | NA | NA | NA | NA | NA | NA | NA |

| Scoliosis | Joint hypermobility or laxity | Autonomic features | Constipation | Immunological issues | Immunological issues detail | Cancer | Age (range) at last HbF meaurement (yrs) | Increased HbF | HbF % | Previous genetic analyses | Additional clinical manifestations |
|---|---|---|---|---|---|---|---|---|---|---|---|
| HP:0002650 | HP:0001382 HP:0001388 | HP:0002270 HP:0000965 HP:0030880 HP:0010832 | HP:0002019 | HP:0002715 | | HP:0002664 | | HP:0011904 | | | |
| yes | no | no | yes | no | | no | NA | NA | NA | CMA, Fragile X: normal | |
| yes | no | no | yes | no | | no | NA | NA | NA | CMA, Fragile X: normal. m | |
| no | NA | no | yes | no | normal IgA, IgM and Ig | no | NA | NA | NA | CMA: normal | |
| no | no | no | no | no | | no | NA | NA | NA | CMA: 143 kb 11p13 duplica | left lacrimal duct stenosis |
| no | yes | yes | no | no | | no | NA | NA | NA | CMA, Fragile X: normal | |
| NA | yes | yes | NA | NA | | no | NA | NA | NA | NA | frequent respiratory infections |
| no | no | no | no | NA | | no | 6-10 yrs | yes | 14.7 | *UBE3A seq, MECP2 seq,* k | |
| NA | NA | NA | NA | NA | | no | NA | NA | NA | CMA, karyotype, serum tra | |
| no | yes | no | no | NA | | no | 6-10 yrs | yes | 10.9 | CMA, Fragile X: normal | |
| no | NA | no | no | no | | no | 11-15 yrs | yes | 5.8 | CMA: 1 Kb 11q22.2 deletio | phimosis, concerns for pubertal dev |
| no | no | no | no | no | | no | 11-15 yrs | yes | 19 | CMA, Fragile X, MECP2 se | |
| no | NA | yes | no | NA | | no | NA | NA | NA | CMA: normal | toe walking |
| yes | no | NA | no | no | | no | 16-20 yrs | yes | 16.5 | CMA, Fragile X: normal | |
| no | no | no | no | no | | no | 6-10 yrs | yes | 14.7 | CMA, tranferrin, *CLCN2, C* | hypothyroidism diagnosed in early |
| yes | no | yes | yes | no | | no | 6-10 yrs | yes | 6.3 | karyotype, 15q11q13 meth | |
| NA | yes | NA | yes | no | | no | NA | yes | 4.1 | NA | persistent thrombocytosis |
| no | no | no | no | NA | | no | NA | NA | NA | CMA, *RAI1* seq: normal | abnormal thrombocyte morphology |
| NA | yes | NA | yes | NA | | no | NA | yes | 10.5 | CMA, Fragile X: normal | |
| NA | NA | NA | NA | NA | | no | NA | NA | NA | CMA, Fragile X: normal | abnormal eye movements (Dent sy |
| no | NA | no | yes | NA | | no | 6-10 yrs | yes | 14.7 | mitochondrial myopathy b | |
| NA | no | NA | NA | NA | | no | NA | NA | NA | CMA, Fragile X, microceph | elevated CK |
| no | no | no | no | NA | | no | NA | NA | NA | CMA, *CNTNAP2, NRXN1* s | |
| no | no | no | no | NA | | no | NA | NA | NA | 15q11q13 methylation, *ME* | |
| NA | NA | NA | NA | NA | | no | NA | NA | NA | Xq28 qPCR, *MECP2* seq, *FL* | |
| no | no | no | no | NA | | no | NA | yes | 35 | CMA, Fragile X, SMA, 15q1 | |
| no | no | no | no | no | | no | 6-10 yrs | yes | 15.3 | CMA, Fragile X: normal | |
| NA | yes | NA | NA | NA | | no | NA | yes | high (n | CMA, Fragile X, congenital | migraines, periodic fevers with aph |
| NA | yes | NA | NA | no | | no | NA | yes | 16.3 | NA | hearing loss |
| NA | no | NA | NA | NA | | no | NA | NA | NA | NA | |
| yes | yes | no | no | NA | | no | NA | yes | 26.3 | CMA, Fragile X: normal | precocious puberty |
| no | no | no | no | no | | no | NA | NA | NA | Fragile X: normal. CMA: 3 | cryptorchydidm, repeatedly elevate |
| no | no | no | no | NA | | no | NA | yes | 16 | CMA, 15q11q12 methylati | right cryptorchidism, clinodactyly o |
| no | no | no | no | no | | no | 1-5 yrs | yes | 15.9 | *ATXN1, ATXN2, ATXN3, C* | |
| NA | yes | NA | NA | NA | | no | NA | NA | NA | 15q11q13 methylation: no | cryptorchidism, umbilicated nipples |
| no | yes | yes | no | no | normal IgA, IgM and Ig | no | 6-10 yrs | yes | 20.8 | CMA, Fragile X, MECP2 se | |
| no | no | NA | NA | no | normal IgA, IgM and Ig | no | 16-20 yrs | yes | 6.6 | CMA: normal | |
| no | no | yes | yes | NA | | no | NA | NA | NA | CMA: Xp22.33 duplication, | |
| no | NA | no | no | yes | hypogammaglobuliner | no | 16-20 yrs | yes | 16 | CMA: normal | lacrimal duct stenosis, meatal sten |
| NA | no | no | NA | NA | | no | NA | NA | NA | NA | bilateral inguinal hernia, Duane and |
| no | no | no | yes | NA | | no | 11-15 yrs | yes | 16 | karyotype, Fragile X, *TSC1/* | hearing loss |
| NA | yes | yes | yes | no | normal IgA, IgM and Ig | no (but osteoc | 6-10 yrs | yes | 25.5 | beta Hb gene analysis, mD | branchial cleft cyst, osteochondrom |
| NA | NA | NA | NA | NA | | no | NA | NA | NA | NA | |
| yes | yes | NA | NA | NA | | no | NA | yes | 3.1 | NA | |
| NA | yes | NA | NA | NA | | no | NA | yes | 8.6 | NA | |
| NA | yes | NA | NA | NA | | no | 13 | yes | 7.5 | karyotype, telomeres, CMA | |
| NA | yes | NA | yes | NA | | no | NA | NA | NA | NA | |
| NA | no | NA | NA | NA | | no | NA | yes | 16 | NA | |
| yes | no | NA | NA | NA | | no | 2.5 | yes | 2.4 | NA | |
| NA | no | NA | NA | NA | | no | NA | NA | NA | NA | |
| NA | no | NA | NA | NA | | no | 7 | yes | 4.4 | metabolic (plasma aminoa | |
| NA | no | NA | NA | NA | | no | NA | NA | NA | NA | |
| NA | no | NA | NA | NA | | no | 7.25 | yes | 12.1 | NA | |
| NA | no | NA | NA | NA | | no | NA | NA | NA | CMA: 4.3 Mb 15q15.3q21.1 | delayed bone age, congenital hip d |
| NA | yes | NA | NA | NA | | no | NA | NA | NA | NA | |
| NA | no | NA | NA | NA | | no | NA | NA | NA | NA | |
| NA | yes | NA | yes | NA | | no | NA | yes | 8.7 | NA | |
| NA | yes | NA | NA | NA | | no | NA | yes | 12.7 | NA | |
| NA | NA | NA | NA | NA | | no | NA | NA | NA | NA | |
| NA | NA | NA | NA | NA | | no | NA | NA | NA | karyotype: normal. CMA: 3 | |
| NA | NA | NA | NA | NA | | NA | NA | NA | NA | NA | |
| NA | NA | NA | NA | NA | | NA | NA | NA | NA | NA | |
| NA | NA | NA | NA | NA | | NA | NA | NA | NA | NA | elevated serum CK |
| NA | NA | NA | NA | NA | | NA | NA | NA | NA | NA | |
| NA | NA | NA | NA | NA | | NA | NA | NA | NA | NA | |
| NA | NA | NA | NA | NA | | NA | NA | NA | NA | NA | |
| NA | NA | NA | NA | NA | | NA | NA | NA | NA | NA | |
| NA | NA | NA | NA | NA | | NA | NA | NA | NA | NA | |
| NA | NA | NA | NA | NA | | NA | NA | NA | NA | NA | |
| NA | NA | NA | NA | NA | | NA | NA | NA | NA | NA | |
| NA | NA | NA | NA | NA | | NA | NA | NA | NA | NA | |
| NA | NA | NA | NA | NA | | NA | NA | NA | NA | NA | |
| NA | NA | NA | NA | NA | | NA | NA | NA | NA | NA | |
| NA | NA | NA | NA | NA | | NA | NA | NA | NA | NA | |
| NA | NA | NA | NA | NA | | NA | NA | NA | NA | NA | |
| NA | NA | NA | NA | NA | | NA | NA | NA | NA | WES: KMT2D (VUS) | |

| Patient ID | Mutation class[1] | Subclass | BCL11A-XL - MANE Select transcript | | | | BCL11A-L | | | | BCL11A-S | | | |
|---|---|---|---|---|---|---|---|---|---|---|---|---|---|---|
| | | | NM_022893.4: | NP_075044.2: | Classification | NMD | NM_018014.4: | NP_060484.2: | Classification | NMD | NM_138559.2: | NP_612569.1: | Classification | NMD |
| **P1** | PTV | PTVa1 | c.156_157insCCTG | p.Met53ProfsTer32 | frameshift | NMDeSP | c.156_157insCCTG | p.Met53ProfsTer32 | frameshift | NMDeSP | c.156_157insCCTG | p.Met53ProfsTer32 | frameshift | NMDeSP |
| **P2** | PTV | PTVa1 | c.12_19dup | p.Gly7AlafsTer9 | frameshift | NMDeSP | c.12_19dup | p.Gly7AlafsTer9 | frameshift | NMDeSP | c.12_19dup | p.Gly7AlafsTer9 | frameshift | NMDeSP |
| **P3** | PTV | PTVa1 | c.53C>A | p.Ser18Ter | stop_gain | NMDeSP | c.53C>A | p.Ser18Ter | stop_gain | NMDeSP | c.53C>A | p.Ser18Ter | stop_gain | NMDeSP |
| **P4** | PTV | PTVa1 | c.148C>T | p.Gln50Ter | stop_gain | NMDeSP | c.148C>T | p.Gln50Ter | stop_gain | NMDeSP | c.148C>T | p.Gln50Ter | stop_gain | NMDeSP |
| **P5** | PTV | PTVa2 | c.156_157insCCTG | p.Met53ProfsTer32 | frameshift | NMD+ | c.156_157insCCTG | p.Met53ProfsTer32 | frameshift | NMD+ | c.156_157insCCTG | p.Met53ProfsTer32 | frameshift | NMD+ |
| **P6** | PTV | PTVa2 | c.263C>A | p.Ser88Ter | stop_gain | NMD+ | c.263C>A | p.Ser88Ter | stop_gain | NMD+ | c.263C>A | p.Ser88Ter | stop_gain | NMD+ |
| **P7** | PTV | PTVa2 | c.286_291delinsA | p.Ser96ThrfsTer13 | frameshift | NMD+ | c.286_291delinsA | p.Ser96ThrfsTer13 | frameshift | NMD+ | c.286_291delinsA | p.Ser96ThrfsTer13 | frameshift | NMD+ |
| **P8** | PTV | PTVa2 | c.295del | p.Val99TrpfsTer29 | frameshift | NMD+ | c.295del | p.Val99TrpfsTer29 | frameshift | NMD+ | c.295del | p.Val99TrpfsTer29 | frameshift | NMD+ |
| **P9** | PTV | PTVa2 | c.363C>A | p.Cys121Ter | stop_gain | NMD+ | c.363C>A | p.Cys121Ter | stop_gain | NMD+ | c.363C>A | p.Cys121Ter | stop_gain | NMD+ |
| **P10** | PTV | PTVa1 | c.502dup | p.Ser168LysfsTer69 | frameshift | NMDeLE | c.502dup | p.Ser168LysfsTer69 | frameshift | NMDeEL | c.502dup | p.Ser168LysfsTer74 | frameshift | NMDeEJ |
| **P11** | PTV | PTVb1 | c.633_643del | p.Gly212ArgfsTer21 | frameshift | NMDeLE | c.633_643del | p.Gly212ArgfsTer21 | frameshift | NMDeEL | c.630+3_630+13del | p.(?) | splice_region_variant | |
| **P12** | PTV | PTVb1 | c.794del | p.Leu265ArgfsTer15 | frameshift | NMDeLE | c.794del | p.Leu265ArgfsTer15 | frameshift | NMDeEL | c.630+164del | p.= | na | |
| **P13** | PTV | PTVb1 | c.794del | p.Leu265ArgfsTer15 | frameshift | NMDeLE | c.794del | p.Leu265ArgfsTer15 | frameshift | NMDeEL | c.630+164del | p.= | na | |
| **P14** | PTV | PTVb1 | c.952A>T | p.Arg318Ter | stop_gain | NMDeLE | c.952A>T | p.Arg318Ter | stop_gain | NMDeEL | c.630+322A>T | p.= | na | |
| **P15** | PTV | PTVb1 | c.1078dup | p.Leu360ProfsTer212 | frameshift | NMDeLE | c.1078dup | p.Leu360ProfsTer212 | frameshift | NMDeEL | c.630+448dup | p.= | na | |
| **P16** | PTV | PTVb1 | c.1078dup | p.Leu360ProfsTer212 | frameshift | NMDeLE | c.1078dup | p.Leu360ProfsTer212 | frameshift | NMDeEL | c.630+448dup | p.= | na | |
| **P17** | PTV | PTVb1 | c.1078dup | p.Leu360ProfsTer212 | frameshift | NMDeLE | c.1078dup | p.Leu360ProfsTer212 | frameshift | NMDeEL | c.630+448dup | p.= | na | |
| **P18** | PTV | PTVb1 | c.1118dup | p.Val374GlyfsTer198 | frameshift | NMDeLE | c.1118dup | p.Val374GlyfsTer198 | frameshift | NMDeEL | c.630+488dup | p.= | na | |
| **P19** | PTV | PTVb1 | c.1133C>G | p.Ser378Ter | stop_gain | NMDeLE | c.1133C>G | p.Ser378Ter | stop_gain | NMDeEL | c.630+503C>G | p.= | na | |
| **P20** | PTV | PTVb2 | c.1287_1288insCACA | p.Lys430HisfsTer143 | frameshift | NMDeLE | c.1287_1288insCACA | p.Lys430HisfsTer143 | frameshift | NMDeEL | c.630+657_630+658insCACA | p.= | na | |
| **P21** | PTV | PTVb2 | c.1411A>T | p.Lys471Ter | stop_gain | NMDeLE | c.1411A>T | p.Lys471Ter | stop_gain | NMDeEL | c.630+781A>T | p.= | na | |
| **P22** | PTV | PTVb2 | c.1459G>T | p.Glu487Ter | stop_gain | NMDeLE | c.1459G>T | p.Glu487Ter | stop_gain | NMDeEL | c.630+829G>T | p.= | na | |
| **P23** | PTV | PTVb2 | c.1459G>T | p.Glu487Ter | stop_gain | NMDeLE | c.1459G>T | p.Glu487Ter | stop_gain | NMDeEL | c.630+829G>T | p.= | na | |
| **P24** | PTV | PTVb2 | c.1601_1631del | p.Val534AlafsTer54 | frameshift | NMDeLE | c.1601_1631del | p.Val534AlafsTer54 | frameshift | NMDeEL | c.630+971_630+1001del | p.= | na | |
| **P25** | PTV | PTVb2 | c.1690del | p.Gln564ArgfsTer34 | frameshift | NMDeLE | c.1690del | p.Gln564ArgfsTer34 | frameshift | NMDeEL | c.630+1060del | p.= | na | |
| **P26** | PTV | PTVb2 | c.1735_1741del | p.Glu579ThrfsTer17 | frameshift | NMDeLE | c.1735_1741del | p.Glu579ThrfsTer17 | frameshift | NMDeEL | c.630+1105_630+1111del | p.= | na | |
| **P27** | PTV | PTVb2 | c.1831G>T | p.Glu611Ter | stop_gain | NMDeLE | c.1831G>T | p.Glu611Ter | stop_gain | NMDeEL | c.630+1201G>T | p.= | na | |
| **P28** | PTV | PTVb2 | c.1847dup | p.Leu617ProfsTer18 | frameshift | NMDeLE | c.1847dup | p.Leu617ProfsTer18 | frameshift | NMDeEL | c.630+1217dup | p.= | na | |
| **P29** | PTV | PTVb2 | c.1967_1968dup | p.Ser657ThrfsTer134 | frameshift | NMDeLE | c.1967_1968dup | p.Ser657ThrfsTer122 | frameshift | NMDeEL | c.630+1337_630+1338dup | p.= | na | |
| **P30** | PTV | PTVb2 | c.2035_2036del | p.Ser679GlnfsTer47 | frameshift | NMDeLE | c.2035_2036del | p.Ser679GlnfsTer47 | frameshift | NMDeEL | c.630+1405_630+1406del | p.= | na | |
| **P31** | PTV | PTVb2 | c.2192_2201dup | p.Ser734ArgfsTer15 | frameshift | NMDeLE | c.2192_2201dup | p.Ser734ArgfsTer23 | frameshift | NMDeEJ | c.630+1562_630+1571dup | p.= | na | |
| **P32** | SPL | SPL | c.55+1G>T | p.(?) | splice_donor_loss | | c.55+1G>T | p.(?) | splice_donor_loss | | c.55+1G>T | p.(?) | splice_donor_loss | |
| **P33** | SPL | SPL | c.56-1G>A | p.(?) | splice_acceptor_loss | | c.56-1G>A | p.(?) | splice_acceptor_loss | | c.56-1G>A | p.(?) | splice_acceptor_loss | |
| **P34** | SPL | SPL | c.385+2T>C | p.(?) | splice_donor_loss | | c.385+2T>C | p.(?) | splice_donor_loss | | c.385+2T>C | p.(?) | splice_donor_loss | |
| **P35** | MISS | MISS | c.139A>C | p.Thr47Pro | missense | | c.139A>C | p.Thr47Pro | missense | | c.139A>C | p.Thr47Pro | missense | |
| **P36** | MISS | MISS | c.143G>T | p.Cys48Phe | missense | | c.143G>T | p.Cys48Phe | missense | | c.143G>T | p.Cys48Phe | missense | |
| **P37** | MISS | MISS | c.563A>G | p.His188Arg | missense | | c.563A>G | p.His188Arg | missense | | c.563A>G | p.His188Arg | missense | |
| **P38** | MISS | MISS | c.2268T>G | p.Asn756Lys | missense | | c.2230+38T>G | p.= | na | | c.630+1638T>G | p.= | na | |
| **Previously reported** | | | | | | | | | | | | | | |
| **Dias_8** | PTV | PTVa2 | c.154C>T | p.Gln52Ter | stop_gain | NMD+ | c.154C>T | p.Gln52Ter | stop_gain | NMD+ | c.154C>T | p.Gln52Ter | stop_gain | NMD+ |
| **Dias_9** | PTV | PTVa2 | c.193G>T | p.Glu65Ter | stop_gain | NMD+ | c.193G>T | p.Glu65Ter | stop_gain | NMD+ | c.193G>T | p.Glu65Ter | stop_gain | NMD+ |
| **Korenke_1** | PTV | PTVa2 | c.271del | p.Glu91ArgfsTer2 | frameshift | NMD+ | c.271del | p.Glu91ArgfsTer2 | frameshift | NMD+ | c.271del | p.Glu91ArgfsTer2 | frameshift | NMD+ |
| **Dias_4 (Decipher 268026)** | PTV | PTVa1 | c.529C>T | p.Gln177Ter | stop_gain | NMDeLE | c.529C>T | p.Gln177Ter | stop_gain | NMDeEL | c.529C>T | p.Gln177Ter | stop_gain | NMD+ |
| **Yoshida_1** | PTV | PTVa1 | c.577del | p.His193MetfsTer3 | frameshift | NMDeLE | c.577del | p.His193MetfsTer3 | frameshift | NMDeEL | c.577del | p.His193MetfsTer3 | frameshift | NMDeEJ |
| **Cai_1** | PTV | PTVb1 | c.644C>G | p.Ser215Ter | stop_gain | NMDeLE | c.644C>G | p.Ser215Ter | stop_gain | NMDeEL | c.630+14C>G | p.= | na | |
| **Dias_11** | PTV | PTVb1 | c.793dup | p.Leu265ProfsTer3 | frameshift | NMDeLE | c.793dup | p.Leu265ProfsTer3 | frameshift | NMDeEL | c.630+163dup | p.= | na | |
| **Soblet_1** | PTV | PTVb2 | c.1343del | p.Pro448ArgfsTer31 | frameshift | NMDeLE | c.1343del | p.Pro448ArgfsTer31 | frameshift | NMDeEL | c.630+713del | p.= | na | |
| **Dias_10** | PTV | PTVb2 | c.1325del | p.Leu442ProfsTer37 | frameshift | NMDeLE | c.1325del | p.Leu442ProfsTer37 | frameshift | NMDeEL | c.630+695del | p.= | na | |
| **Wessels_1** | PTV | PTVb2 | c.1453G>T | p.Glu485Ter | stop_gain | NMDeLE | c.1453G>T | p.Glu485Ter | stop_gain | NMDeEL | c.630+823G>T | p.= | na | |
| **Dias_6 (Decipher 280953)** | PTV | PTVb2 | c.1540_1544dup | p.Phe515LeufsTer5 | frameshift | NMDeLE | c.1540_1544dup | p.Phe515LeufsTer5 | frameshift | NMDeEL | c.630+910_630+914dup | p.= | na | |
| **Dias_7** | PTV | PTVb2 | c.1775_1776insTGGCTCAGCGG | p.Glu593GlyfsTer9 | frameshift | NMDeLE | c.1775_1776insTGGCTCAGCGG | p.Glu593GlyfsTer9 | frameshift | NMDeEL | c.630+1145_630+1146insTGGCTCAGCGG | p.= | na | |
| **Cai_2** | PTV | PTVb2 | c.1826del | p.Pro609ArgfsTer21 | frameshift | NMDeLE | c.1826del | p.Pro609ArgfsTer21 | frameshift | NMDeEL | c.630+1196del | p.= | na | |
| **Dias_3** | MISS | MISS | c.198C>A | p.His66Gln | missense | | c.198C>A | p.His66Gln | missense | | c.198C>A | p.His66Gln | missense | |
| **Yoshida_2** | MISS | MISS | c.2351A>C | p.Lys784Thr | missense | | c.2230+121A>C | p.= | na | | c.630+1721A>C | p.= | na | |
| **Publicly accessibile databases** | | | | | | | | | | | | | | |
| **Decipher 291669** | PTV | PTVa1 | c.16C>T | p.Gln6Ter | stop_gain | NMD+ | c.16C>T | p.Gln6Ter | stop_gain | NMD+ | c.16C>T | p.Gln6Ter | stop_gain | NMD+ |
| **VCV000987092.1** | PTV[2] | PTVa1[2] | HGVSc=c.55_55+1insT | p.Pro19LeufsTer5 | frameshift + splice_do | NMD+ | HGVSc=c.55_55+1insT | p.Pro19LeufsTer5 | frameshift + splice_dc | NMD+ | HGVSc=c.55_55+1insT | p.Pro19LeufsTer5 | frameshift + splice_d | NMD+ |
| **VCV000987335.1** | PTV | PTVa2 | c.370C>T | p.Gln124Ter | stop_gain | NMD+ | c.370C>T | p.Gln124Ter | stop_gain | NMD+ | c.370C>T | p.Gln124Ter | stop_gain | NMD+ |
| **Decipher 261208** | PTV | PTVa1 | c.596T>G | p.Leu199Ter | stop_gain | NMDeLE | c.596T>G | p.Leu199Ter | stop_gain | NMDeEL | c.596T>G | p.Leu199Ter | stop_gain | NMDeEJ |
| **Decipher 400685** | PTV | PTVa1 | c.599_602del | p.Glu200AlafsTer7 | frameshift | NMDeLE | c.599_602del | p.Glu200AlafsTer7 | frameshift | NMDeEL | c.599_602del | p.Glu200AlafsTer7 | frameshift | NMDeEJ |
| **VCV000986112.1** | PTV | PTVb1 | c.794del | p.Leu265ArgfsTer15 | frameshift | NMDeLE | c.794del | p.Leu265ArgfsTer15 | frameshift | NMDeEL | c.630+164del | p.= | na | |
| **VCV000987079.1** | PTV | PTVb1 | c.1078del | p.Leu360SerfsTer61 | frameshift | NMDeLE | c.1078del | p.Leu360SerfsTer61 | frameshift | NMDeEL | c.630+448del | p.= | na | |
| **VCV000986806.1** | PTV | PTVb2 | c.1555_1556del | p.Leu519GlyfsTer52 | frameshift | NMDeLE | c.1555_1556del | p.Leu519GlyfsTer52 | frameshift | NMDeEL | c.630+925_630+926del | p.= | na | |
| **VCV000985399.1** | PTV | PTVb2 | c.2194dup | p.Arg732LysfsTer14 | frameshift | NMDeLE | c.2194dup | p.Arg732LysfsTer22 | frameshift | NMDeEJ | c.630+1564dup | p.= | na | |
| **VCV000985925.1** | SPL | SPL | c.55+1G>A | p.(?) | splice_donor_loss | | c.55+1G>A | p.(?) | splice_donor_loss | | c.55+1G>A | p.(?) | splice_donor_loss | |
| **VCV000987091.1** | SPL | SPL | c.55+1_55+2insCCCAA | p.(?) | splice_donor | | c.55+1_55+2insCCCAA | p.(?) | splice_donor | | c.55+1_55+2insCCCAA | p.(?) | splice_donor | |
| **VCV000987090.1** | SPL | SPL | c.55+5del | p.(?) | splice_donor_loss | | c.55+5del | p.(?) | splice_donor_loss | | c.55+5del | p.(?) | splice_donor_loss | |
| **VCV000987093.1** | MISS[3] | MISS[3] | c.55C>T | p.Pro19Ser | missense + splice | | c.55C>T | p.Pro19Ser | missense + splice | | c.55C>T | p.Pro19Ser | missense + splice | |

***Supplementary Table S2.*** Sequence variants in *BCL11A* included in the full clinical cohort.

For all cases ClinVar and Decipher IDs are indicated; IDs starting with "VCV" are ClinVar references. MANE Select transcript: matched annotation between NCBI and EBI. 1For mutation class description, see Methods. 2frameshift + splice site disruption (SpliceAI, donor_gain and donor_loss). 3missense and potential splice site disruption (SpliceAI). NMD+, expected to elicit nonsense mediated decay; NMDe, expected to escape NMD (significantly truncated protein); SP, start proximal (≤150 nt from start codon); EL, long exon (exon length ≥407 nt); EJ, exon juntion (≤ 50 nt from last exon junction); LE, last exon.

| CNVs | | |
|---|---|---|
| **Patient ID** | **Microdeletion** | **Description** |
| **P39** | [GRCh38] 2p16.1(60508596_60555113)x1 | gene disruption: deletion of UTR > intron 2; encompasses lncRNA LOC102724142 |
| **P40** | [GRCh38] 2p16.1(60533641_60561918)x1 | gene disruption: deletion of upstream NCR, 5'UTR > intron 2 |
| **P41** | [GRCh38] 2p16.1(59833747_60655840)x1 | whole gene deletion of BCL11A and includes MIR4432, MIR4432HG and lncRNA LOC102724142 |
| **P42** | [GRCh38] 2p16.1(60466611_60575172)x1 | gene disruption: deletion of upstream NCR, 5'UTR > intron 3 |
| **Previously reported** | | |
| **Balci_1** | [GRCh38] 2p16.1(59731285_60607163)x1 | whole gene deletion of BCL11A; includes MIR4432, MIR4432HG and lncRNA LOC102724142 |
| **Peter_1** | [GRCh38] 2p16.1(60462164_60603356)x1 | gene disruption deletion of upstream NCR, 5'UTR > exon4 |
| **Publicly accessible databases** | | |
| **Decipher_300584** | [GRCh38] 2p16.1(60452463_60629834)x1 | whole gene deletion of BCL11A and UTR region with no coding or non-protein-coding genes |
| **Decipher_300585** | [GRCh38] 2p16.1(60452463_60629834)x1 | whole gene deletion of BCL11A and UTR region with no coding or non-protein-coding genes |
| **Decipher_305786** | [GRCh38] 2p16.1(60214168_60469017)x1 | gene disruption from intron 2 > 3'UTR and includes MIR4432, MIR4432HG and lncRNA LOC102724142 |

***Supplementary Table S3.*** Copy number variants in *BCL11A* included in the full clinical cohort.

| Reference | Patient ID | Sex | Age (yrs) | CNV | Congenital malformations | Detail of congenital malformations | brain MRI | Detail of brain MRI | Posterior fossa abn | Brain stem abn | Corpus callosum abn | Cortical malformation |
|---|---|---|---|---|---|---|---|---|---|---|---|---|
| Rajcan-Separovic et al., 2007 (refined in Liu et al., 2011) | **Rajcan-Separovic_1** | F | 8 | 6.1 Mb 2p15p16.1 microdeletion, de novo, involving BCL11A and 37 additional genes (+ paternally inherited 1.2 Mb Xp22.31 deletion) | yes | multicystic kidney | abnormal | bilateral perisylvian cortical dysplasia, optic nerve hypoplasia | no | no | no | yes |
| Rajcan-Separovic et al., 2007 (refined in Liu et al., 2011) | **Rajcan-Separovic_2** | M | 6 | 7.9 Mb 2p15p16.1 microdeletion, de novo, involving BCL11A and 51 additional genes | yes | hypoplastic genitalia | abnormal | mild hypoplasia of inferior cerebellar vermis, enlarged 4th ventricle, small anterior pituitary and small pons, cortical dysplasia, optic nerve hypoplasia | yes | yes | no | yes |
| de Leeuw et al., 2008 | **de Leeuw_1** | M | 32 | 3.9 Mb mosaic 2p15p16.1 microdeletion, de novo, involving BCL11A and 9 additional genes | yes | inguinal hernia, atrophic right testis | NA | NA | NA | NA | NA | NA |
| Liang et al., 2009 | **Liang_1** | F | 4.5 | 3.2 Mb 2p15p16.1 microdeletion, de novo, involving BCL11A and 10 additional genes | no | | normal | normal | no | no | no | no |
| Felix et al., 2010 | **Felix_1** | F | 4 | 3.35 Mb 2p15p16.1 microdeletion, de novo, involving BCL11A and 11 additional genes | no | | normal | normal | no | no | no | no |
| Hucthagowder et al., 2012 | **Hucthagowder_1** | F | 2 | 2.5 Mb 2p15p16.1 microdeletion, de novo, involving BCL11A and 8 additional genes | yes | multiple renal cysts, giant congenital choledochal cysts | abnormal | simplified gyral pattern, hypoplasia of corpus callosum | no | no | yes | yes |
| Piccione et al., 20121 (and Funnell et al., 2015) | **Piccione_1** | F | 2 | 642 Kb 2p16.1 microdeletion, de novo, involving BCL11A and 4 additional genes (+ paternally inherited 930 Kb 6q12 deletion) | yes | hypoplastic labia minora | normal | normal | no | no | no | no |
| Florisson et al., 2013 | **Florisson_1** | M | 4 | 6.8 Mb 2p15p16.1 microdeletion, de novo, involving BCL11A and 34 additional genes | yes | craniosynostosis | abnormal | simplified gyral pattern in the supratentorial region, hypoplasia of corpus callosum, small aspect of the cerebellum and pons | yes | yes | yes | yes |
| Florisson et al., 2013 | **Florisson_2** | F | 13 | 6.9 Mb 2p15p16.1 microdeletion, de novo, involving BCL11A and 42 additional genes | yes | craniosynostosis | NA | NA | NA | NA | NA | NA |
| Hancarova et al., 2013 (and Basak et al., 2015 patient 1) | **Hancarova_1** | F | 11 | 450 Kb 2p16.1 microdeletion, de novo, involving BCL11A and 3 additional genes | no | | normal | normal | no | no | no | no |
| Jorgez et al., 2014 | **Jorgez_4** | M | 1.75 | 6.31 Mb 2p16.1 microdeletion, de novo, involving BCL11A and several additional genes | yes | mesocardia, renal ectopia, micropenis | abnormal | cerebral atrophy, prominent ventricles, enlarged cysterna magna | yes | no | no | no |
| Basak et al., 2015 | **Basak_2** | NA | NA | 1 Mb 2p16.1 microdeletion, de novo, involving BCL11A and 2 additional genes | NA | NA | normal | normal | no | no | no | no |
| Ottolini et al., 2015 | **Ottolini_1** | F | 11 | 1.07 Mb 2p15p16.1 microdeletion, de novo, involving BCL11A and 7 additional genes | yes | ventricular septal defect | NA | NA | NA | NA | NA | NA |
| Bagheri et al., 2016 | **Bagheri_1** | M | 4 | 9.6 Mb 2p16.1 microdeletion, de novo, involving BCL11A and several additional genes | no | | abnormal | cortica dysplasia | NA | NA | NA | yes |
| Bagheri et al., 2016 | **Bagheri_3** | M | 3 | 5.4 Mb 2p16.1 microdeletion, de novo, involving BCL11A and several additional genes | no | | normal | normal | no | no | no | no |
| Bagheri et al., 2016 | **Bagheri_4** | F | 1.8 | 971 Kb 2p16.1 microdeletion, de novo, involving BCL11A and 10 additional genes | yes | unspecified cardiac anomaly, unspecified genital anomaly | NA | NA | NA | NA | NA | NA |
| Codipilly et al., 2017 | **Codipilly_1** | M | 42 | 4.9 Mb 2p16.1 microdeletion, de novo, involving BCL11A and 29 additional genes | yes | cryptorchidism | abnormal | asymmetric lateral ventricles, prominent cisterna magna | yes | no | no | no |
| Levy et al., 2017 | **Levy_3** | M | 7 | 440 Kb 2p16.1 microdeletion, de novo, involving BCL11A and 1 additional gene | no | | abnormal | enlargement of lateral ventricles, cortical and subcortical atrophy, mild cerebellar atrophy especially in the inferior part | yes | no | no | no |
| Shimbo et al., 2017 | **Shimbo_1** | F | 3 | 3.24 Mb 2p15p16.1 microdeletion, de novo, involving BCL11A andseveral additional genes | no | | abnormal | mild hypoplasia of the pons, hypoplasia of the cerebellum, delayed myelination | yes | yes | no | no |
| Shimbo et al., 2017 | **Shimbo_2** | F | 1.8 | 5.04 Mb 2p15p16.1 microdeletion, de novo, involving BCL11A and several additional genes | yes | anal atresia | abnormal | hypoplasia of the pons, hypoplasia of the corpus callosum, cerebellar hypoplasia | yes | yes | yes | no |
| Shimbo et al., 2017 | **Shimbo_4** | M | 4 | 1.12 Mb 2p16.1 microdeletion, de novo, involving BCL11A and 2 additional genes | no | | abnormal | hypoplasia of the pons and cerebellum | yes | yes | no | no |

***Supplementary Table S5.*** Summary of congenital malformations and brain MRI findings in previously reported individuals with large deletions encompassing additional genes.

| ID | Mutation class[1] | PTV subclass | BCL11A-XL - MANE Select transcript | | | | BCL11A-L | | | | BCL11A-S | | | |
|---|---|---|---|---|---|---|---|---|---|---|---|---|---|---|
| | | | NM_022893.4: | NP_075044.2: | Classification | NMD | NM_018014.4: | NP_060484.2: | Classification | NMD | NM_138559.2: | NP_612569.1: | Classification | NMD |
| rs267599415 | PTV | PTVa2 | c.743C>A | p.Ser248Ter | stop_gain | NMD+ | c.743C>A | p.Ser248Ter | stop_gain | NMD+ | c.630+113C>A | p.= | intron_variant | |
| rs746438740 | PTV | PTVb | c.2185_2197del | .Gly729ProfsTer5 | frameshift | NMDe | c.2185_2197del | .Gly729ProfsTer4 | frameshift | NMDe | 0+1555_630+156 | p.= | intron_variant | |
| rs1315265032 | PTV | PTVb[1] | c.2400C>G | p.Tyr800Ter | stop_gain | NMDe | c.2230+170C>G | p.= | intron_variant | | c.630+1770C>G | p.= | intron_variant | |
| rs772387514 | PTV | PTVb[1] | c.2436C>G | p.Tyr812Ter | stop_gain | NMDe | c.2230+206C>G | p.= | intron_variant | | c.630+1806C>G | p.= | intron_variant | |
| rs771597942 | PTV | PTVb[1] | c.2476C>T | p.Arg826Ter | stop_gain | NMDe | c.2230+246C>T | p.= | intron_variant | | c.630+1846C>T | p.= | intron_variant | |
| rs754847260 | PTV | PTVb[2] | na | na | ıstream_gene_variant | | c.2265del | .Arg756GlufsTer2 | frameshift | NMDe | c.665del | .Arg222LysfsTer1 | frameshift | NMDe |
| rs1265922527 | PTV | PTVb[3] | na | na | ıstream_gene_variant | | c.2303C>T | p.Ser768Leu | missense | | c.703C>T | p.Arg235Ter | stop_gain | NMDe |
| rs1359171807 | MISS[4] | | c.2384T>A | p.Val795Glu | missense | | c.2230+154T>A | p.= | intron_variant | | c.630+1754T>A | p.= | intron_variant | |

***Supplementary Table S7.*** Loss of function sequence variants in *BCL11A* in GnomAD.

[1]Predicted to escape NMD in BCL11A-XL and not affect BCL11A-L or BCL11A-S

[2]Predicted to escape NMD in BCL11A-L and BCL11A-S and not affect BCL11A-XL; frameshift in stop gain in NM_001363864.1 and XM_011532910.1 isoforms

[3]Predicted to escape NMD in BCL11A-S, cause a missense change in BCL11A-L, and not affect BCL11A-XL; stop gain in NM_001363864.1 and XM_011532910.1 isoforms

[4]Classified as LoF based on effect of NM_001363864.1 and XM_011532910.1 isoforms: splice_donor_variant
